# Prenatal exposure to SARS-CoV-2, early relational health, and child socio-emotional functioning in the first 6 months

**DOI:** 10.64898/2026.03.12.26346895

**Authors:** Andréane Lavallée, Jennifer M. Warmingham, Jill B. Owen, Ruiyang L. Xu, Imaal Ahmed, Ginger D. Atwood, Margaret H Kyle, Maha Hussain, Vitoria Chaves, Elena Arduin, Marissa R. Lanoff, Sabrina P. Hyman, Lerzan Z. Coskun, Nicole Shearman, Jenna E. Russo, Sharon Ettinger, Esther A. Greeman, Danielle E. Serota, Mary L. Bence, Violet Hott, Yunzhe Hu, Georgia Kurman, Mayerly Lara, Helen Tzul Lopez, Isabelle Mollicone, Rupa Ravi, Cynthia Rodriguez, Grace C. Smotrich, Aoife Lawless, Perla Ontiveros-Ángel, Austen Curtin, Judy Austin, Morgan R. Firestein, Lauren C. Shuffrey, Cristina R. Fernández, Ashley N. Battarbee, Ann Bruno, Fatimah S. Dawood, Panagiotis Maniatis, Tyler C. Morrill, Gabriella Newes-Adeyi, Lawrence Reichle, Vera Semenova, Alan T. Tita, Michael Varner, Kristina Wielgosz, Rachel Marsh, Paul Curtin, Melissa S. Stockwell, Catherine Monk, Dani Dumitriu

**Author notes:** **Corresponding Authors:** Dani Dumitriu, MD, PhD, Center for Early Relational Health, 3 Columbus Circle, FL 11, New York, NY 10019.

## Abstract

**Importance:** Parent/caregiver-infant early relational health (ERH) is known to play a critical role in the promotion of socio-emotional functioning and wellbeing across the life course. The negative impact of the COVID-19 pandemic on maternal mental health and secondarily on ERH and child socio-emotional functioning is clear. However, the direct impact of maternal viral exposure during pregnancy on ERH has not been investigated.

**Objective:** The goal of this study was to determine the impact of prenatal SARS-CoV-2 exposure on ERH and infant socio-emotional functioning in the first 6 months of life.

**Design:** Mothers with and without SARS-CoV-2 exposure during pregnancy who gave birth from 02/2020 to 09/2021 were enrolled from 05/2020 to 09/2021 in one of two parallel prospective studies (the COVID-19 Mother Baby Outcomes [COMBO] Initiative or the Respiratory Syndrome Coronavirus 2 in Pregnancy and Infancy [ESPI] COMBO sub-study). Mothers reported on their health and the socioemotional functioning of their infant via online surveys (REDCap) at enrollment, 1, 2, 4, and 6 months. At 4 to 6 months, dyads were invited to participate in a video-based, remote assessment of ERH.

**Participants:** 884 mother-infant dyads from three U.S. States (Alabama, New York, and Utah).

**Exposure:** Prenatal SARS-CoV-2.

**Main Outcomes and Measures:** Maternal-reported ERH (parental stress, parenting confidence and bonding) and observer-based ERH (video-coded quality of maternal caregiving behaviors and mother-infant emotional connection). Infant socio-emotional development assessed using the 6-month Ages and Stages Questionnaire: Socio-Emotional 2^nd^ Edition (ASQ:SE-2).

**Results:** 316 (36%) mothers had a positive prenatal SARS-CoV-2 exposure. Prenatal SARS-CoV-2 exposure was associated with an adjusted estimate of ∼5% reduction (incidence rate ratio=0.95, 95% confidence interval [0.90, 1.00], p=0.03) in observed maternal caregiving quality, after accounting for postnatal maternal mental health and sociodemographic factors. We found no evidence of effect on other ERH constructs or infant socio-emotional functioning.

**Conclusions and Relevance:** In this large prospective cohort study, prenatal SARS-CoV-2 was associated with a small decrement in caregiving quality, but not other ERH constructs or infant socioemotional functioning. These findings should be interpreted as hypothesis generating and will require replication in independent studies.

**Key Points:**

- **Question**: Is SARS-CoV-2 exposure during pregnancy associated with maternal-reported and observer-based measures of early relational health (ERH) and infant socio-emotional functioning at 4-6 months postpartum?
- **Findings**: Prenatal SARS-CoV-2 exposure was associated with a ∼5% reduction in observed quality of maternal caregiving after accounting for postnatal mental health symptomatology and sociodemographic factors.
- **Meaning**: A small reduction in maternal caregiving quality, but not other ERH constructs (parental stress, parenting confidence, bonding and emotional connection) or infant socio-emotional functioning, was associated with prenatal SARS-CoV-2 exposure. Results should be interpreted as hypothesis generating and will require replication in independent studies.

## Background

Parent-infant positive and nurturing early relationships – hereafter Early Relational Health (ERH) – buffer the negative effects of childhood adversity^1-7^ and play a critical role in the promotion of child socio-emotional functioning.^8-10^ The COVID-19 pandemic caused a worldwide upsurge in psychological distress, affecting pregnant and postpartum women.^11-23^ This rise in perinatal psychopathology has been associated with disruptions in *maternal-reported* measures of ERH, including bonding,^24-30^ sensitive parenting,^31^ and parenting self-efficacy,^32^ and child socio-emotional functioning.^33-36^ In contrast, only one small study (n=43 dyads) investigated associations between COVID-19 pandemic stressors and emotional availability, and *observer-based* measures of ERH.^37^ Substantial changes to the social and family environment (e.g., social distancing, mask wearing)^38^ and perinatal care (e.g., canceled prenatal visits, separation at birth)^39^ may have increased the burden on parents and disrupted the transition to parenthood.

The impact of the pandemic on maternal mental health and secondarily on ERH and child socioemotional functioning has been extensively documented. Far less research has investigated the effects of prenatal viral exposure on these outcomes. Only one study reported a non-significant association between prenatal SARS-CoV-2 and postnatal *maternal-reported* bonding.^40^ Three studies found no evidence of prenatal SARS-CoV-2-related delays in socioemotional functioning at 3,^41^ 12,^42,43^ and 24^42^ months of age.

Despite evidence of robust fetal protection against vertical transmission of SARS-CoV-2,^44-46^ maternal immune activation (MIA) may still have consequences for mother and infant wellbeing and behavior postnatally.^47-50^ MIA has been associated with altered infant behavior in the first year of life^51,52^, which in turn could impact ERH.^53^ SARS-CoV-2 may affect maternal neurobiology via inflammatory pathways, which can manifest postnatally as neurological symptoms relevant to ERH, such anosmia,^54,55^ fatigue, and brain fog.^56^ Relative to non-exposed pregnant women, those with SARS-CoV-2 exposure during pregnancy are also at higher risk for complications, including hospitalization, caesarean delivery, preterm birth, severe perinatal morbidity and mortality indices,^57-62^ and acute stress responses to childbirth.^62^

Our primary objectives were to determine the impact of prenatal SARS-CoV-2 exposure on ERH and infant socio-emotional functioning at 4-6 months postpartum. To our knowledge, this study is the first to comprehensively evaluate the impact of prenatal viral exposure on maternal-reported and observer-based measures of ERH and infant socio-emotional functioning in a large sample of mother-infant dyads in the context of the COVID-19 pandemic. As the effects of SARS-CoV-2 during pregnancy on ERH remain largely unknown, we examined ERH outcomes across multiple domains (i.e., parental stress, parenting confidence, bonding, caregiving behaviors, and emotional connection), using a combination of *maternal-reported* and *observer-based* measures. Our team developed a remote-adapted, observer-based assessment of ERH, employing videoconferencing technology to record in participants’ homes. This approach allowed flexibility for participants, increased ecological validity, reduced travel burden, and allowed us to reach underserved communities.^63-66^

## Methods

### Study Design and Participants

Mother-infant dyads with and without prenatal SARS-CoV-2 exposure were enrolled into two prospective cohort studies: the COMBO Initiative (formerly the COVID-19 Mother Baby Outcomes cohort) at Columbia University’s Irving Medical Center (CUIMC) or the Centers for Disease Control’s (CDC) Epidemiology of Severe Acute Respiratory Syndrome Coronavirus 2 in Pregnancy and Infancy (ESPI) Network Community Cohort. A subset of ESPI participants from three participating sites (CUIMC, University of Alabama in Birmingham [UAB], and University of Utah [UU]) were enrolled in an ESPI COMBO sub-study mirroring assessments in the COMBO Initiative. Enrollment occurred from 05/2020 to 09/2021. Recruitment, enrollment strategies, and determination of SARS-CoV-2 status for COMBO and ESPI COMBO have been previously described^67^ (**eMethods1**).

CUIMC implemented universal nasopharyngeal polymerase chain reaction (PCR) clinical testing on 03/22/2020, and universal serological testing for SARS-CoV-2 antibodies on 07/20/2020. In the COMBO Initiative, infants born before 11/01/2020 were considered exposed if mother had a positive SARS-CoV-2 PCR and/or serological test during pregnancy or at delivery in her electronic health record (EHR). After 11/01/2020, infants were considered exposed only if mother had a positive PCR or antigen test during pregnancy. Infants were considered unexposed if all PCR and serological tests available in the EHR for the mother were negative, which was estimated to have a 0.67% false-negative rate.^67^

In the ESPI COMBO cohort, infants were classified as exposed if their mother had SARS-CoV-2 infection detected from study surveillance samples (weekly swab collection and molecular testing) or had evidence of seroconversion from study enrollment through the end of the pregnancy.

This study was reviewed and approved by the CUIMC institutional review board (IRB) (^§^ See 45 C.F.R. part 46.114; 21 C.F.R. part 56.114) and all participants provided written informed e-consent, including the use of video/photographic footage. Participants received financial compensation for all study procedures. Participants were also provided with appropriate referrals for services for mothers (e.g. counseling) and/or for infants requiring care (e.g. our Pediatric clinic).

Dyads included in this analysis were born from February 2020 through September 2021. Mothers were invited to complete surveys via electronic self-administration (REDCap) at enrollment, 1, 2, 4, and 6 months postpartum, and were invited to participate in an ERH Zoom-based assessment at 4 to 6 months. Assessments were completed in English or Spanish. Mothers were not required to contribute longitudinal data to remain enrolled. Study design allowed for catch-up if survey time-points were missed.

### Outcome Assessments

#### Maternal-reported ERH

##### Parental Stress Scale

At 6 months postpartum, mothers completed the Parental Stress Scale, an 18-item questionnaire scored on a 5-point Likert scale assessing feelings about the parenting role.^68^ Items were summed to form a total score, with higher scores indicating higher stress.

##### Karitane Parenting Confidence Scale

Mothers completed the Karitane Parenting Confidence Scale (KPCS) at 1, 2, or 4 months postpartum in the COMBO Initiative, and 4 or 6 months in the ESPI COMBO sub-study. The KPCS is a 15-item questionnaire scored on 4-point Likert scale, developed to assess perceived parenting self-efficacy.^69^ Items were summed, with higher total scores indicating higher parenting confidence.

##### Postpartum Bonding Questionnaire –Revised

At 4 months postpartum, mothers completed the Postpartum Bonding Questionnaire-Revised (PBQ-R), a 14-item questionnaire on a 6-point Likert scale, previously validated by our team.^70^ Items were summed to form a total score, with higher scores indicating stronger bonding.

#### Remote Observer-Based Assessment of ERH

At 4 to 6 months postpartum, dyads were invited to participate in a ∼45-min recorded video call. Dyads engaged in 3 minutes of face-to-face interaction with the infant in the mother’s lap as they normally would, followed by a 10-min double Still-Face paradigm (i.e., mothers interacting with their infant as they normally would, interrupted by 2-min periods of either Still-Face [mother focusing her gaze above infant in a neutral way and not responding to infant] or interacting with mask-wearing), concluded by 2 minutes of cuddling. Finally, mothers were asked to change their infant’s diaper. The visit ended with a semi-structured interview focusing on the dyad’s experience during the COVID-19 pandemic. For reliability, 20% of videos were independently coded by two trained coders on all coding schemes described below.

##### Maternal Caregiving Behaviors

The diaper change procedure was coded for quality of maternal caregiving behaviors (MCB) using a coding scheme adapted from Ainsworth.^71^ Subscales included acceptance vs. rejection, sensitivity vs. insensitivity, consideration vs. intrusiveness, and the quality of the mother’s physical contact, vocal contact, and positive affect. Each subscale was scored on a 9-point Likert scale. Scores were combined to produce a total score. Higher scores represent sensitive, non-intrusive, and gentle handling with delight and positive affect during diapering.

##### Emotional Connection

The 3-min face-to-face interaction with infant in mother’s lap was coded using an adaptation of the Welch Emotional Connection Screen,^72^ a brief assessment of dyadic ERH where mutuality in approach and flow in face-to-face interactive behaviors were each coded on a 9-point Likert scale.^72^ Higher scores indicate stronger mother-infant emotional connection.

#### Infant Socio-Emotional Functioning

##### Ages and Stages Questionnaire: Socio-Emotional – 2^nd^ Edition

Infant socio-emotional functioning was assessed using the 6-month Ages and Stages Questionnaire: Socio-Emotional 2^nd^ Edition (ASQ:SE-2).^73^ The ASQ:SE-2 is a 23-item maternal report using a 3-point Likert scale, including an item that allows caregivers to indicate if the behavior is of concern (+5 points). Items were summed, with a higher total score indicating higher risk for socioemotional delays. A score between 30 to 45 suggests that some behaviors are of concern and should be monitored; scores ≥45 suggests a need for further assessment.

#### Covariates

##### Maternal Mental Health in the Postpartum Period

Mothers completed surveys assessing mental health symptoms from 4-6 months postpartum (**eMethods 2** for full description of surveys). The Patient Health Questionnaire^74,75^ (PHQ-9) was used to assess depressive symptoms. The State-Trait Anxiety Inventory-State^76^ (STAI-S) assessed state anxiety symptoms. The Posttraumatic Stress Disorder Checklist for DSM‐5^77,78^ (PCL‐5) was modified to measure posttraumatic stress disorder symptoms related to the COVID-19 pandemic. The Perceived Stress Scale^79^ (PSS) was used to assess frequency of subjective stress experiences. The Brief Symptom Inventory^78^ (BSI) Anxiety and Somatization subscales were used to assess general anxiety and somatic-related complaints associated with anxiety, respectively.

##### Demographic data

Sociodemographic variables (maternal age, maternal education, maternal race, maternal ethnicity, parity, gestational age/preterm status) were assessed using demographic surveys collected at each survey timepoint and/or abstracted from EHRs.

### Statistical Analysis

Sociodemographic characteristics of dyads with and without SARS-CoV-2 exposure were compared using non-parametric Kruskal-Wallis tests (continuous variables) or Chi-squared/Fisher’s exact tests (categorical variables). We assessed inter-rater reliability on the observational video-coded outcome data using intraclass correlations (ICC). To test the effect of prenatal SARS-CoV-2 on six outcomes (parental stress, parenting confidence, bonding, maternal caregiving behaviors, emotional connection, and child socio-emotional functioning), we fit Poisson regression models with a log link function (generalized linear models [GLM]) to account for non-negative integer outcome measures. In case of overdispersion, we fit negative binomial regression models. We implemented unadjusted and adjusted models for all outcomes. Adjusted models’ covariates included: maternal age at delivery, race (White, Black/African American, other, declined), ethnicity (non-Hispanic, Hispanic, declined), insurance (commercial, Medicaid), mode of delivery (vaginal, caesarean-section), parity (multiparous, primiparous), maternal mental health symptomatology in the first 6 months postpartum, infant sex (male, female), infant age at time of assessment (corrected for prematurity), and study site (CUIMC, UU, UAB). Percentages of infants meeting the cut-off indicating risk for socioemotional delays (score ≥30) per ASQ:SE-2 developers^73^ were computed and compared using GLM with binomial distribution to determine whether those differed by exposure groups. Analyses included only participants with complete data due to listwise deletion. Statistical analyses were conducted in R version 4.4.2.^80^ Significance level was set at α<0.05.

The covariate for maternal mental health symptomatology in the first 6 months postpartum was operationalized as a factor score generated from a confirmatory factor analysis (Mplus v. 8.6.42) of the six maternal mental health questionnaires described above (*see* **eMethods 3** and **eTable 1**).

To visualize the relationship between outcomes and SARS-CoV-2 exposure status while accounting for confounding variables, we used a residual-based adjustment approach. First, we fit covariate-only models for each outcome that included all covariates except SARS-CoV-2 status. We then calculated the residuals from these models—representing the variation in each outcome not explained by the covariates— and added them to the outcome mean to re-center the data. These adjusted outcomes were plotted against SARS-CoV-2 status, allowing us to visualize the association between SARS-CoV-2 exposure and each outcome after controlling for other variables in the model.

In sensitivity analyses, we ran the adjusted models described above in two subsamples. First, we removed dyads who had a confirmed exposure to SARS-CoV-2 before birth, but for which timing of exposure could not be established with certainty (before vs. during pregnancy). Second, we removed dyads with infant date of birth in 02/2020, before the official onset of the COVID-19 pandemic in the U.S.

## Results

### Participant Characteristics

Overall, 884 mother-infant dyads with infants born from 02/2020 through 09/2021 were enrolled in either COMBO or ESPI COMBO (see **Figure 1**) and completed surveys and/or video visits from birth to 6 months. Of those, 316 (36%) mothers had a prenatal SARS-CoV-2 exposure identified by molecular testing of respiratory samples or serologic testing (see **Table 1** for sample characteristics by exposure groups and **eTable 2** for characteristics by study sites). Overall, infection severity in the COMBO and ESPI COMBO cohorts was mild in approximately 75% of women, with the remaining ∼25% being asymptomatic.^67^ Exposure groups were unbalanced; SARS-CoV-2 exposed mothers were younger, more represented by women identifying as Multiracial or Hispanic, and more frequently used Medicaid insurance and gave birth before term. Of 316 mothers with positive SARS-CoV-2 tests, 53 had an unknown timing of exposure that could have been before or after conception (see **eTable 3** for sample characteristics by three exposure groups: negative, positive, positive-timing unknown). The distributions of all outcomes by SARS-CoV-2 exposure are in **Figure 2**.

**Figure 1.**
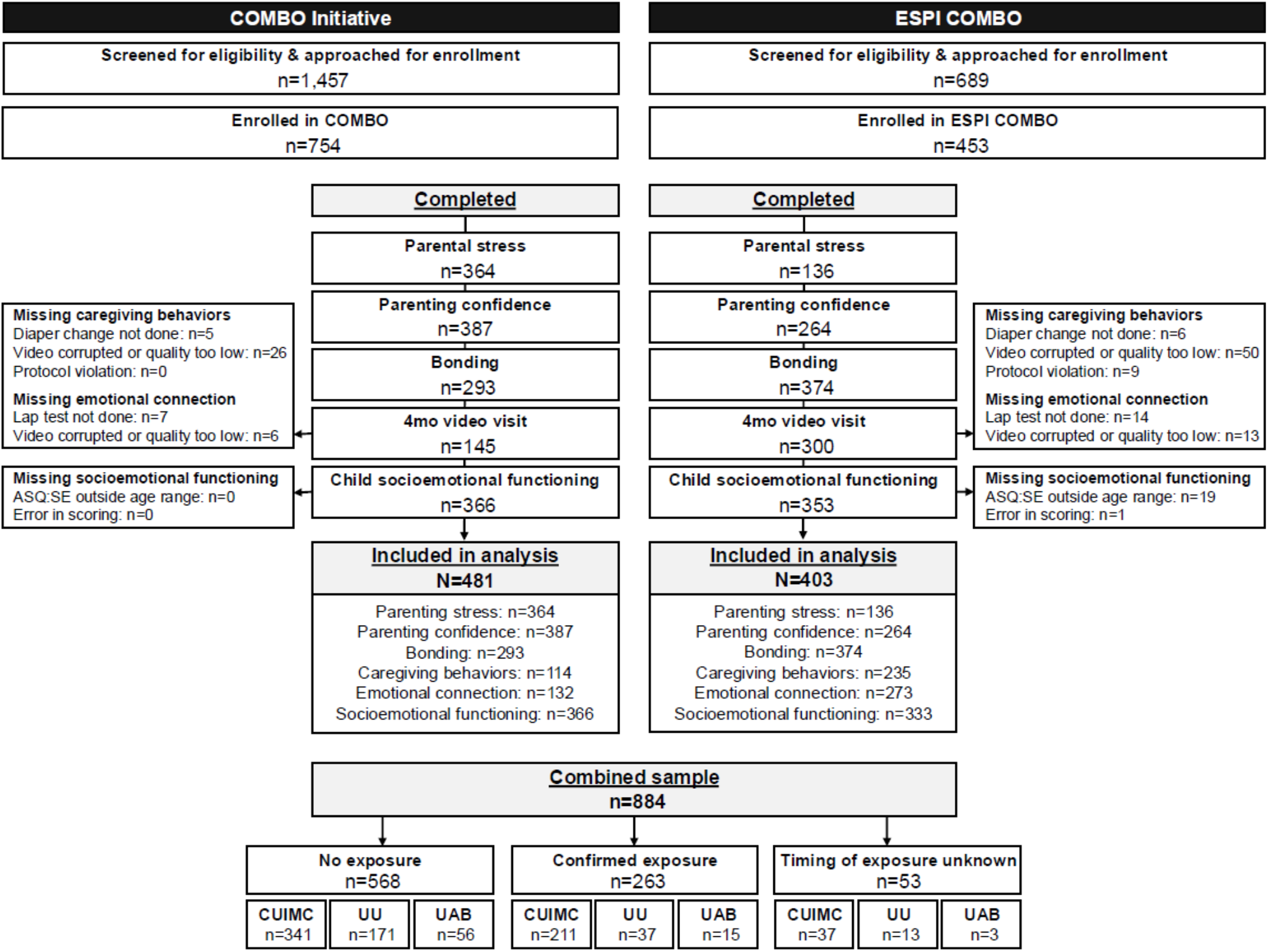
COMBO Initiative and ESPI COMBO participant flow chart.

**Table 1.** Characteristics of mother-infant dyads by SARS-CoV-2 exposure status.

|  | Overall | Negative | Positive | p-value |
| --- | --- | --- | --- | --- |
|  | N=884 | n=568 | n=316 |  |
| **Maternal age at delivery** |  |  |  |  |
| Mean (SD) | 32.2 (5.4) | 32.6 (5.3) | 31.6 (5.5) | **0.027** |
| Range | 17.0-47.0 | 18.3-45.3 | 17.0-47.0 |  |
| **Maternal race** |  |  |  | **<0.001** |
| White | 456 (52.1) | 334 (59.3) | 122 (39.1) |  |
| Other/More than one category | 180 (20.6) | 86 (15.3) | 94 (30.1) |  |
| Black/African American | 100 (11.4) | 59 (10.5) | 41 (13.1) |  |
| Asian | 43 (4.9) | 36 (6.4) | 7 (2.2) |  |
| Native Hawaiian/ Pacific Islander | 7 (0.8) | 3 (0.5) | 4 (1.3) |  |
| Native American/Alaska Native | 4 (0.5) | 3 (0.5) | 1 (0.3) |  |
| Declined | 84 (9.6) | 42 (7.5) | 42 (13.5) |  |
| Missing | 1 (0.1) | 0 (0.0) | 1 (0.3) |  |
| **Maternal ethnicity** |  |  |  | **<0.001** |
| Not Hispanic | 508 (58.1) | 380 (67.5) | 128 (41.0) |  |
| Hispanic | 336 (38.4) | 164 (29.1) | 172 (55.1) |  |
| Declined | 31 (3.5) | 19 (3.4) | 12 (3.8) |  |
| Missing | 0 (0.0) | 0 (0.0) | 0 (0.0) |  |
| **Insurance** |  |  |  | **<0.001** |
| Private | 552 (63.1) | 399 (70.9) | 153 (49.0) |  |
| Medicaid | 321 (36.7) | 163 (29.0) | 158 (50.6) |  |
| Missing | 2 (0.2) | 1 (0.2) | 1 (0.3) |  |
| **Mode of delivery** |  |  |  | 0.79 |
| Vaginal | 559 (63.9) | 355 (63.1) | 204 (65.4) |  |
| C-Section | 316 (36.1) | 208 (36.9) | 108 (34.6) |  |
| Missing | 0 (0.0) | 0 (0.0) | 0 (0.0) |  |
| **Parity** |  |  |  | 0.061 |
| Multiparous | 472 (53.9) | 287 (51.0) | 185 (59.3) |  |
| Primiparous | 403 (46.1) | 276 (49.0) | 127 (40.7) |  |
| Missing | 0 (0.0) | 0 (0.0) | 0 (0.0) |  |
| **Infant sex** |  |  |  | 0.955 |
| Male | 476 (53.8) | 308 (54.2) | 168 (53.2) |  |
| Female | 408 (46.2) | 260 (45.8) | 148 (46.8) |  |
| Missing | 0 (0.0) | 0 (0.0) | 0 (0.0) |  |
| **Twin status** |  |  |  | 0.839 |
| Singleton | 866 (99.0) | 558 (99.1) | 308 (98.7) |  |
| Twin pairs | 9 (1.0) | 5 (0.9) | 4 (1.3) |  |
| Missing | 0 (0.0) | 0 (0.0) | 0 (0.0) |  |
| **Prematurity status** |  |  |  | **0.046** |
| Not Preterm | 801 (90.6) | 525 (92.4) | 276 (87.3) |  |
| Preterm | 83 (9.4) | 43 (7.6) | 40 (12.7) |  |
| Missing | 0 (0.0) | 0 (0.0) | 0 (0.0) |  |
| **Infant corrected age at survey completion** |  |  |  |  |
| Mean (SD) |  |  |  |  |
| Parental stress | 6.0 (0.4) | 6.0 (0.4) | 6.0 (0.4) | 0.216 |
| Parenting confidence | 3.0 (2.0) | 3.0 (2.0) | 2.9 (1.9) | 0.678 |
| Bonding | 4.7 (1.6) | 4.7 (1.6) | 4.5 (1.4) | 0.442 |
| Infant socio-emotional development | 6.2 (0.9) | 6.2 (0.9) | 6.2 (0.9) | 0.846 |
| **Infant corrected age at video visit** |  |  |  |  |
| Mean (SD) | 5.4 (2.2) | 5.6 (2.3) | 5.0 (1.9) | **0.037** |

**Figure 2.**
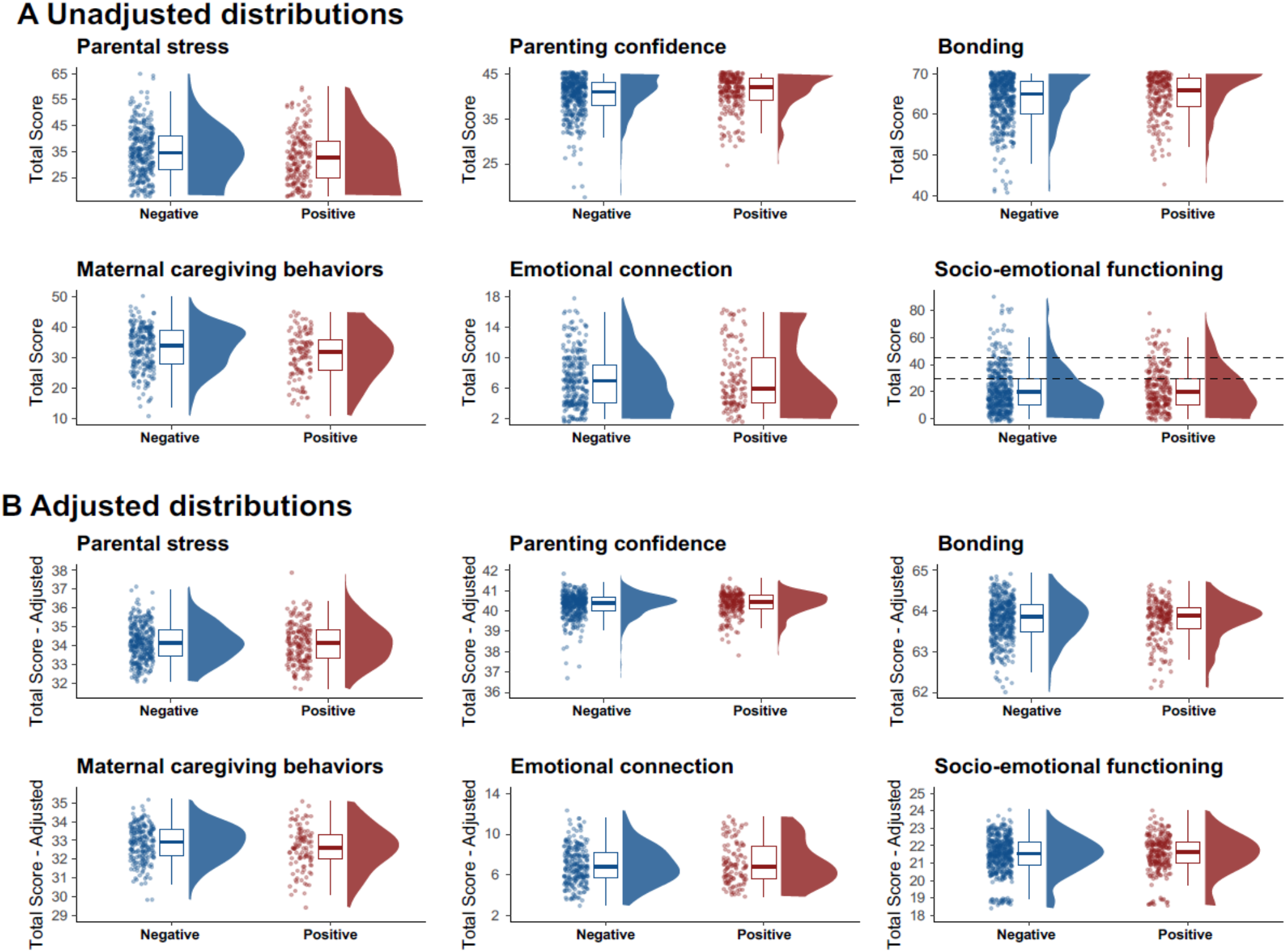
Outcome data distribution by prenatal SARS-CoV-2 exposure status. Negative: no documental prenatal SARS-CoV-2 exposure. Positive: confirmed prenatal SARS-CoV-2 exposure (including timing unknown). For COMBO Initiative participants, positive status was determined by universal nasopharyngeal polymerase chain reaction (PCR) testing and universal serological testing during pregnancy and at birth. For ESPI COMBO participants, positive status was determined with weekly nasopharyngeal collection and molecular testing, and by periodic serum collection to evaluate for seroconversion. Dotted lines on socio-emotional functioning plot represent cut-off score of 30 above which socio-emotional development should be monitored, and score of 45 above which socio-emotional development should be assessed by a professional per ASQ:SE-2 developers. **Panel A** represents raw distributions. **Panel B** represents outcomes after covariate adjustment (see Methods).

### Prenatal Exposure to SARS-CoV-2 and Maternal-Report of Early Relational Health

Prenatal SARS-CoV-2 exposure was associated with a 5.4% decrease in parental stress in the unadjusted model (n=500, incidence rate ratio [IRR]=0.95, 95% confidence interval (95% CI) [0.90, 0.99], p=0.03, **Figure 3A**), but not in the adjusted model (n=499, IRR=1.00, 95%CI [0.95, 1.05], p=0.91, **Figure 3A** and **eTable4**). SARS-CoV-2 exposure was not associated with parenting confidence (unadjusted: n=651, IRR=1.02, 95%CI [0.99, 1.04], p=0.21, **Figure 3B**; adjusted: n=621, IRR=1.01, 95%CI [0.98, 1.03], p=0.61, **Figure 3B** and **eTable5**) or bonding (unadjusted: n=667, IRR=1.02, 95%CI [1.00, 1.04], p=0.08, **Figure 3C**; adjusted: n=659, IRR=1.00, 95%CI [0.98, 1.03], p=0.76, **Figure 3C** and **eTable 6**). Sensitivity analyses confirm these results (**eTable 7** to **eTable 12**).

**Figure 3.**
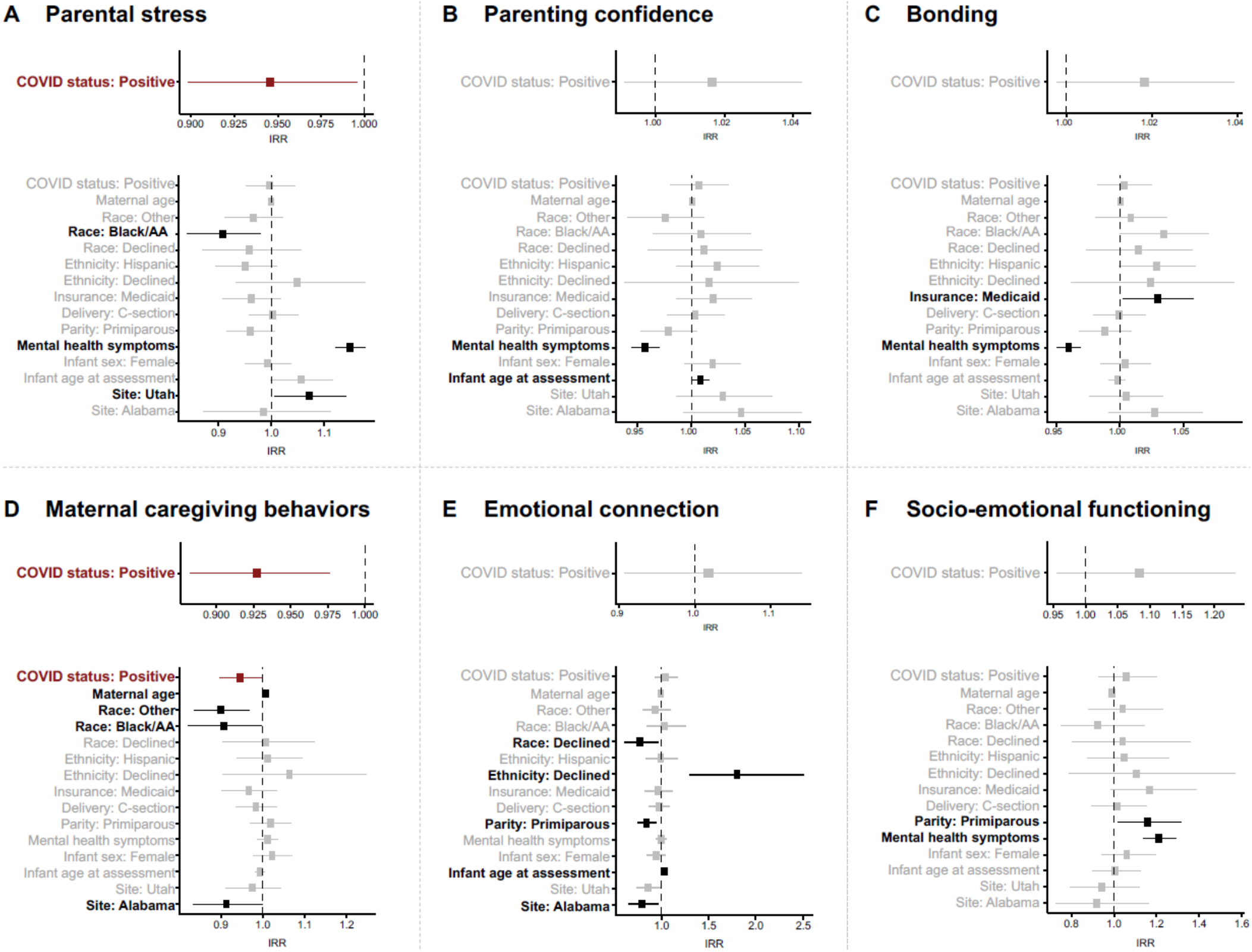
Generalized linear model estimates of the effect of prenatal SARS-CoV-2 exposure on ERH and infant socio-emotional functioning. Significant predictors are in bolded in red for SARS-CoV-2 status and black for other model predictors. **Panel A:** Parental stress reported by mothers on the Parental stress scall. Higher score indicated more stress. **Panel B:** Parenting stress reported by mothers on the Karitane Parenting Confidence Scale. Higher score indicated more confidence. **Panel C:** Bonding reported by mothers on the Postpartum Bonding Questionnaire – Revised. Higher score indicates stronger bonding. **Panel D:** Maternal caregiving behaviors video coded from a diaper change interaction using an adaptation of Ainsworth’s maternal sensitivity scales. Higher score indicates higher caregiving quality. **Panel E:** Emotional connection video coded from a mother-infant face-to-face interaction with infant in mother’s lap using an adaptation of the Welch Emotional Connection Screen. Higher score indicates stronger emotional connection. Panel F: Infant socioemotional functioning reported by mothers on the 6-month Ages and Stages Questionnaire: Socio-Emotional-2^nd^ Edition. **Note. AA:** African American. C-section: cesarean section. IRR: incidence rate ratio.

### Prenatal Exposure to SARS-CoV-2 and Observer-Based Measures of Early Relational Health

Coders achieved reliability with ICCs >0.85 for each behavior from the MCB rating system, and >0.95 on the emotional connection screen. Prenatal SARS-CoV-2 exposure was associated with a 7.2% decrease in quality of maternal caregiving behaviors in the unadjusted model (n=349, IRR=0.93, 95%CI [0.88, 0.98], p=0.004, **Figure 3D**). This association remained significant after adjusting with covariates (n=317, IRR=0.95, 95%CI [0.90, 1.00], p=0.03, **Figure 3D** and **eTable13**).

Prenatal SARS-CoV-2 exposure was not associated with emotional connection (unadjusted: n=405, IRR=1.02, 95%CI [0.91, 1.14], p=0.76, **Figure 3E**; adjusted: n=364, IRR=1.05, 95%CI [0.93, 1.17], p=0.45, **Figure 3E** and **eTable14**). Sensitivity analyses generally confirm these results (**eTable15** to **eTable18**), but the association with maternal caregiving behaviors was marginally significant in the subset without mothers with uncertain timing of exposure (n=291, IRR=0.95, 95%CI [0.90, 1.00], p=0.05, **eTable15)**.

### Prenatal Exposure to SARS-CoV-2 and Socio-emotional Development

Exposure to prenatal SARS-CoV-2 was not associated with ASQ:SE-2 scores (unadjusted: n=698, IRR=1.08, 95%CI [0.95, 1.23], p=0.22, **Figure 3F**; adjusted: n=696, IRR=1.06, 95%CI [0.93, 1.20], p=0.41, **Figure 3F** and **eTable19**). There was no significant effect on the overall risk of socio-emotional delays (unadjusted: n=698, IRR=1.33, 95%CI [0.81, 2.17], p=0.26; adjusted: n=696, IRR=1.19, 95%CI [0.67, 2.08], p=0.55, **eTable20**). Sensitivity analyses supported these results (**eTable21** to **eTable24**).

## Discussion

This study is the first to examine associations between prenatal SARS-CoV-2 exposure, maternal-reported and observer-based measures of ERH, and child socio-emotional development in the first 6 months of life. At 4 to 6 months postpartum, we found a ∼5% reduction in observed caregiving quality in mothers exposed to prenatal SARS-CoV-2 compared to unexposed mothers, which remained significant after accounting for the effect of postnatal mental health symptomatology and sociodemographic factors.

The theoretical mechanisms by which prenatal exposure could affect maternal caregiving quality is unclear. Viral exposure during pregnancy may directly alter maternal neurobiology, and in turn, disrupt postnatal caregiving. MIA models in rodents and nonhuman primates suggest that exposure to viral mimetics like poly(I:C) or LPS during pregnancy induces persistent caregiving deficits including reduced licking/grooming, delayed pup retrieval, and poor nest quality.^81,82^ These impairments are associated with key substrates for maternal motivation and bonding, including neuroinflammatory changes in the medial preoptic area (mPOA) and disruptions in oxytocin receptor signalling.^83^ Elevated IL-6 –present in SARS-CoV-2 infected individuals^84^ –and other pro-inflammatory cytokines may impair maternal neural plasticity by disrupting oxytocinergic signalling and caregiving-related circuitry. Consistent with this, gestational immunogenic insults can bias neural processing of both threat and reward, ultimately manifesting as reduced motivation for infant caregiving.^85^

Alternatively, differences in risk of exposure to SARS-CoV-2 (e.g., household size, occupation) could have overlapping mechanisms associated with maternal caregiving behaviors. The unknown effects of prenatal SARS-CoV-2 on the developing foetus could also have shaped maternal postnatal behavior, although unlikely given the absence of association between maternal postnatal mental health and caregiving behaviors in this sample.

Importantly, the magnitude of difference in quality in caregiving behavior between the exposed and unexposed group was small. We found no evidence that prenatal SARS-CoV-2 exposure affects other ERH constructs, including parental stress, parenting confidence, bonding and emotional connection. Given the number of hypotheses tested, there is an increased risk of type I error. Our findings should therefore be interpreted as hypothesis generating and will require replication in independent samples.

Aligning with previously published data showing that maternal prenatal SARS-CoV-2 exposure is not associated with early signs of delays in child development,^86-90^ we found no association with infant socio-emotional functioning at 6 months. Also, consistent with research from before^91-95^ and during the COVID-19 pandemic,^24-28,31^ we found that greater maternal postpartum mental health symptoms were associated with higher parental stress, lower parenting confidence and bonding, and more infant socio-emotional problem behavior. Critically, these effects did not extend to *observer-based* ERH constructs (caregiving behaviors and emotional connection), suggesting that mothers who experienced more mental health symptoms in the postpartum period *subjectively* experienced less closeness with their infants.

## Limitations

As previously described,^67,87^ unmeasured confounding variables, such as psychosocial factors, could affect ERH and socio-emotional functioning. Most mothers in this sample experienced asymptomatic or mild infection. Therefore, results cannot be generalized to infants prenatally exposed to moderate to severe infections. In the COMBO Initiative, a potential for misclassification of exposure status is possible given that this study was not a prospective surveillance cohort. Measurement error related to maternal-reported measures and potential social desirability bias during performative video sessions may have influenced results. Separately, the smaller proportion of mothers who completed the Zoom-based assessment compared to surveys could explain discrepancies between maternal-reported and observer-based outcomes. Finally, follow-up of these dyads is needed to monitor the association between prenatal SARS-CoV-2 exposure and longer-term ERH and child socioemotional functioning, especially given the documented association between maternal caregiving behaviors and the emergence of child neurodevelopment.^96^

## Conclusion

Prenatal SARS-CoV-2 was associated with a small decrement in caregiving quality, but not other ERH constructs or infant socio-emotional functioning. Maternal mental health symptoms were associated with a decrement in *maternal-reported* but not *observer-based* ERH measures. These results suggest there may be a need for coordinated, long-term follow-up of the health and wellbeing of pandemic-born children and their parents^49^ to advance our understanding of the risk and resilience processes resulting from the COVID-19 pandemic. Results should be replicated in independent studies, and suggest long-term follow-up of the COVID-19 generation given the well-established role of early-life sensitive caregiving on later child attachment^97^ and behavior.^96^ Longitudinal assessment of multimodal neurobiological and longitudinal approaches, including neuroimaging, and neuroendocrine profiling could explore candidate pathways by which prenatal viral exposure may disrupt maternal caregiving circuitry. Such work would advance our understanding of how viral exposures shape ERH and offspring socioemotional development and to identify biological targets for precision intervention to support positive caregiving.

## Data Availability

All data produced in the present study are available upon reasonable request to the authors.

## Acknowledgements

Dr. Lavallée had full access to all study data and takes responsibility for the integrity of the data and the accuracy of the data analysis. Written informed consent was obtained from study participants and staff for publication of photographs.

## Contributions

Concept and design: AL, JMW, JBO, MHK, MRF, LCS, CRF, RM, MS, PC, CM, DD.

Acquisition, analysis, or interpretation of data: AL, JMW, JBO, RLX, IA, GA, MHK, MH, VC, GA, EA, MRL, NS, SE, EAG, AL, DES, MLB, VH, YH, GK, MK, ML, HTL, IEM, RR, CR, GCS, AC, JA, ANB, AB, FSD, SG, PM, TM, GNA, LR, VS, ATT, MV, KW.

Drafting of the manuscript: AL, JMW, SPH, LC, JER, POA, DD.

Critical revision of the manuscript for important intellectual content: JMW, JBO, RLX, IA, GA, MHK, MH, VC, EA, MRL, SPH, LC, NS, JER, SE, EAG, AL, DES, MLB, VH, YH, GK, MK, ML, HTL, IEM, RR, CR, GCS, POA, AC, JA, MRF, LCS, CRF, ANB, AB, FSD, SG, PM, TM, GNA, LR, VS, ATT, MV, KW, RM, MS, PC, CM, DD.

Obtained funding: AL, AT, MV, MS, RM, CM, DD.

Administrative, technical, or material support: PC

Supervision: PC, CM, DD.

## Data Sharing

The datasets generated during and/or analyzed during the current study are available from the corresponding author on reasonable request.

## Conflict of Interest Disclosures

None to disclose.

## Funding/Support

This study was funded by the US Centers for Disease Control and Prevention through contract 75D30120C08150 with Abt Associates (Drs Gyamfi-Bannerman, Mourad, Stockwell, and Dumitriu), Einhorn Collaborative (Dumitriu), grant R01MH126531 from the National Institute of Mental Health (Drs Marsh, Monk, and Dumitriu), fellowships from the Canadian Institutes of Health Research and *Fonds de Recherche en Santé du Québec* (Lavallée) and 1K99HD115784-01 from Eunice Kennedy Shriver National Institute of Child and Human Development (Lavallée).

## Role of the Funder/Sponsor

Authors affiliated with the U.S. Centers for Disease Control and Prevention were involved in the design and conduct of the study; the analysis and interpretation of the data; the preparation, review, and approval of the manuscript; and decision to submit the manuscript for publication. These authors were not directly involved in the collection and management of the data. The National Institutes of Mental Health and the Eunice Kennedy Shriver National Institute of Child Health and Human Development had no role in the design and conduct of the study; collection, management, analysis, and interpretation of the data; preparation, review, or approval of the manuscript; and decision to submit the manuscript for publication.

## Disclaimer

The findings and conclusions in this article are those of the authors and do not necessarily represent the views of the US Centers for Disease Control and Prevention.

## Notes

### Competing Interest Statement

The authors have declared no competing interest.

### Author Declarations

IRB of Columbia University gave ethical approval for this work.

