## Supplemental files for "Prenatal exposure to SARS-CoV-2, early relational health, and child socio-emotional functioning in the first 6 months"

### eMethods1. Determination of SARS-CoV-2 infection status

**COVID-19 Mother-Baby Outcomes (COMBO) Initiative**
Beginning May 2020, the COMBO Initiative recruited women who received prenatal care and delivered at the Columbia University Irving Medical Center (CUIMC)-affiliated NewYork-Presbyterian (NYP) Morgan Stanley Children's Hospital (MSCH) or NYP Allen Pavilion Hospital. Mothers could also self-refer.

***SARS-CoV-2 Positive Group:*** Universal nasopharyngeal PCR testing and universal serological testing for SARS-CoV-2 antibodies was implemented by CUIMC for all delivering mothers between March 22, 2020, and July 20, 2020, respectively. For infants born before November 1, 2020, prenatal exposure was determined if the mother had a positive SARS-CoV- 2 PCR and/or serology test documented in the electronic health record during pregnancy or at delivery. After November 1, 2020, positive SARS-CoV-2 status was determined by a positive PCR or antigen test during pregnancy or a positive serology test and documentation of COVID-19 symptoms in the medical record (determined via electronic chart review).

***SARS-CoV-2 Negative Group:*** Following delivery, each infant born to a mother with a confirmed SARS-CoV-2 infection during pregnancy was matched to one to three infants without documented exposure to a maternal SARS-CoV-2 infection during pregnancy. For infants born before July 20, 2020, the unexposed classification required the absence of PCR positivity in the EHR, no self-reported PCR positivity, absence of COVID-19 symptoms recorded in the EHR and no self-reported COVID-19 symptoms. After July 20, 2020, the unexposed group classification required the absence of PCR positivity and/or a negative serology. Exposed and unexposed groups were matched based on sex, gestational age at birth, mode of delivery, and birthdate within a two-week window.

**ESPI COMBO**
COMBO sub-study. The Epidemiology of Severe Acute Respiratory Syndrome Coronavirus-2 in Pregnancy and Infancy (ESPI) study was a prospective SARS-CoV-2 infection surveillance study that enrolled pregnant women from three academic medical centers in the U.S. Women were eligible if they were pregnant and at <28 weeks gestation; aged 18–50 years; willing to self-collect and mail mid-turbinate nasal swab specimens and respond to weekly surveillance contacts; willing to have up to three blood draws during pregnancy/postpartum; willing to have data collected from their infants' medical records at delivery; and able to speak and read either English or Spanish. Women were ineligible if they were enrolled in a COVID-19 or influenza vaccine clinical trial or intended to enroll in a trial during the current pregnancy. Participants self-collected and submitted weekly mid-turbinate nasal swabs for SARS-CoV-2 reverse transcription polymerase chain reaction (RT-PCR) testing, completed weekly questionnaires about illness symptoms, and submitted additional mid-turbinate swabs when experiencing COVID-19–like symptoms. Serum was collected at enrollment, at the end of the second trimester and at end of pregnancy for testing for SARS-CoV-2 antibodies.

Classification of perinatal SARS-CoV-2 status in this study was based on a combination of maternal self-report of COVID-19 prior to ESPI enrollment, PCR testing and symptom report during the ESPI phase, and serological testing during the ESPI phase.

Serological testing in ESPI was conducted from sera collected up to three times during pregnancy (at enrollment, at end of second trimester, and at end of pregnancy). All participants had serological testing performed at the end of pregnancy. Samples were processed at the CDC using Luminex xMAP-SARS-CoV-2 Multi Antigen Assay. The assay tests 3 antigens: S1, RBD, and nucleocapsid (N) proteins. The N protein is unique to natural infection. S1 and RBD antibodies form in response to both natural infection and vaccination. Considering all three antibodies allows for differentiation between positive testing in response to vaccination versus natural infection. The ESPI sample contained individuals positive for S1 and/or RBD (negative for N), who had never been vaccinated, which was attributed to quicker waning of antibodies against N than of antibodies against S1 and RBD.

The following classification considering self-report, PCR results, and antibody positivity was created:

1. **SARS-CoV-2 not detected**
    a) No self-reported COVID-19 and
    b) Absent positive PCR from ESPI enrollment to delivery and
    c) Negative S1, RBD, N on all serological testing in ESPI and

d) Serology testing was obtained at the end of pregnancy and was negative

2. **SARS-CoV-2 detected: Pre-pregnancy infection with no in utero exposure**
    a) Self-reported COVID-19 infection with a specific date prior to calculated conception date and
    b) Serology consistent with prior infection
        i. Positive N at entry into ESPI or
        ii. Positive RBD/S1 at entry into ESPI and no history of vaccination
3. **SARS-CoV-2 detected: Unknown timing, possible in utero exposure**
    a) No self-report of COVID-19 history, but:
        i. Positive N at entry into ESPI or
        ii. Positive RBD/S1 and no history of vaccination
4. **SARS-CoV-2 detected during pregnancy**
    a) PCR+ from ESPI enrollment to delivery or
    b) Serological conversion from ESPI enrollment to delivery that cannot be explained by vaccination:
        i. Serological conversion with N pos irrespective of vaccine and symptoms or
        ii. Serological conversion with RBD/S1 prior to vaccine irrespective of symptoms

### eMethods2. Maternal Mental Health Questionnaires

- **The Patient Health Questionnaire (PHQ-9)**[1,2] is a 9-item self-report measure that assesses depression symptom severity. Mothers completed this survey online. Mothers indicated how frequently they experienced each of the 9 items (e.g., “Feeling down, depressed, or hopeless”) in the previous two weeks. Response options were 0 (not at all), 1 (several days), 2 (more than half the days), or 3 (nearly every day). Individual item responses were summed to yield a total score (0-27), with higher scores indicating greater severity depression symptoms.

- **The State-Trait Anxiety Inventory-State (STAI-S)**[3] is a 20-item self-report measure that assesses state anxiety. This measure has been used to measure fluctuations in anxiety symptoms in the peripartum period in prior studies.[4] Mothers rated the intensity of their state anxiety (sample items: “I am tense; I am worried” and “I feel calm; I feel secure.”). Response options included 1 (Not at all), 2 (Somewhat), 3 (Moderately so), or 4 (Very much so). A total score was computed such that higher scores indicated greater state anxiety.

- **The Posttraumatic Stress Disorder (PTSD) Checklist for DSM-5 (PCL-5)**[5,6] is a 20-item self-report measure to assess posttraumatic stress disorder symptoms, based on the DSM-5 PTSD criteria. Mothers completed this survey remotely via online survey link. The prompt for this measure was modified in the present study to request that mothers report on severity of PTSD symptoms related to the COVID-19 pandemic. Mothers reported on the severity of each of the 20 symptoms (e.g., “Having strong physical reactions when you thought of COVID-19 (for example, heart pounding, trouble breathing, sweating)” and “Having strong negative feelings such as fear, horror, anger, guilt, or shame related to COVID-19?”) from 0 (Not at all) to 4 (Extremely) in the last month. The PCL-5 has been successfully modified in prior studies to assess COVID-19-related PTSD symptoms.[7]

- **The Perceived Stress Scale (PSS)**[8] is a 14-item self-report measure to assess subjective stress. Items include questions such as “In the last month, how often have you felt nervous or “stressed”?” and “In the last month, how often have you found that you could not cope with all the things that you have had to do?” Mothers responded to each item on a scale from 0 (Never) to 4 (Very often), indicating the frequency of this symptom in the last month. Items were summed to yield a total score, with higher scores indicating greater stress.

- **The Brief Symptom Inventory (BSI)**[6] Anxiety (6 items) and Somaticization (6 items) subscales were completed by mothers via online survey. Mothers reported on their distress related to each item (e.g., “Spells of terror or panic” and “Feeling tense or keyed up”) in the last 7 days on a scale of 1 (Not at all) to 5 (Extremely). Each subscale was scored with higher scores indicating greater symptom presentation.

### eMethods3. Factor Scores for Maternal Postnatal Mental Health Symptomatology

This appendix describes the methods and results of confirmatory factor analyses used to create factor scores for maternal mental health and COVID-19 social impact used in main analyses.

**Methods – Confirmatory Factor Analysis**

Two confirmatory factor analyses (CFAs) were conducted in Mplus (v8.6[9]) to establish latent factors for maternal mental health and COVID-19 social impact. For maternal mental health, six continuous indicators were used. For each of the following, higher scores indicated greater incidence of symptoms in that domain: PHQ-9 score (depression), PCL-5 symptoms related to COVID-19 stress (PTSD symptoms), STAI-state score (anxiety), Perceived Stress Scale score (perceived stress), and the Somatization and Anxiety subscales of the Brief Symptom Index.

For the maternal mental health CFA, Maximum likelihood (ML) estimation was used. Model fit was determined by fit indices including Chi-Squared ($\chi^2$) value, the comparative fit index (CFI), standardized root mean squared residual (SRMR), and root mean squared error of approximation (RMSEA). Goodness of model fit was determined by nonsignificant values of the $\chi 2$ statistic, values of the CFI greater than .95, SRMR values less than .08, and RMSEA values smaller than .05.[10,11]

**Results – Confirmatory Factor Analysis**

The total N for the maternal mental health CFA was N=888. Specific Ns for each measure are listed in **eTable1**. Full information maximum likelihood was used to estimate missing data and use all possible data collected across measures to inform the latent factor.[12] After initial CFA showed poor fit, a correlated residual was specified between the two BSI indicators. Fit was adequate ($\chi^2(8)=62.57$, $p<.001$; RMSEA=.088; CFI=.96, SRMR=.042). Loadings were positive and significant and ranged from .52-.84 (see **eTable1**), indicating that higher scores on the latent factor indicate greater mental health symptoms. The correlated residual between BSI Anxiety and BSI Somatization was also significant ($\beta$=.52, SE=.030, $p<.001$).

***Creation of Factor Scores.*** After finalizing both CFA solutions, factor scores estimates were saved and exported to R to be used as a predictor of early relational health outcome measures. Mplus produces factor score estimates based on the CFA models using the maximum posterior distribution of the factor.[13] This method yields a continuous estimate for each latent factor. Factor scores were standardized (z-scored) after they were exported prior to use in analyses.

### eTable1. Maternal Mental Health Confirmatory Factor Analysis Loadings

| | N | λ | SE | *p*-value |
|---|---|---|---|---|
| PHQ-9 | 744 | .84 | .02 | <.001 |
| STAI-S | 626 | .74 | .03 | <.001 |
| BSI Somatization | 736 | .52 | .0 | <.001 |
| BSI Anxiety | 737 | .59 | .030 | <.001 |
| Perceived Stress Scale | 738 | .73 | .02 | <.001 |
| PCL-5 PTSD Symptoms | 624 | .57 | .03 | <.001 |
| **Total N** | **888** | | | |

Notes: PHQ-9: Patient Health Questionnaire-9 (depression symptoms); STAI-S: State Trait Anxiety Inventory (state score); BSI: Brief Symptom Inventory; PCL-5: PTSD Checklist for DSM-5 (COVID version); λ = standardized factor loading; SE= standard error of factor loading.

**eTable2. Characteristics of mother-infant dyads by study site**

| | Overall | New York | Utah | Alabama | *p*-value |
|---|---|---|---|---|---|
| | **N=884** | **n=589** | **n=221** | **n=74** | |
| **Maternal age at delivery** | | | | | |
| Mean (SD) | 32.2 (5.4) | 32.4 (5.6) | 31.7 (5.1) | 32.2 (4.0) | 0.346 |
| **Maternal race** | | | | | **<0.001** |
| White | 456 (52.1) | 215 (36.8) | 193 (88.5) | 48 (65.8) | |
| Other/More than one category | 180 (20.6) | 163 (27.9) | 14 (6.4) | 3 (4.1) | |
| Black/African American | 100 (11.4) | 78 (13.4) | 1 (0.5) | 21 (28.8) | |
| Asian | 43 (4.9) | 36 (6.2) | 6 (2.8) | 1 (1.4) | |
| Native Hawaiian/ Pacific Islander | 7 (0.8) | 7 (1.2) | 0 (0.0) | 0 (0.0) | |
| Native American/ Alaska Native | 4 (0.5) | 3 (0.5) | 1 (0.5) | 0 (0.0) | |
| Declined | 84 (9.6) | 81 (13.9) | 3 (1.4) | 0 (0.0) | |
| Missing | 1 (0.1) | 1 (0.2) | 0 (0.0) | 0 (0.0) | |
| **Maternal ethnicity** | | | | | **<0.001** |
| Not Hispanic | 508 (58.1) | 239 (40.9) | 199 (91.3) | 70 (95.9) | |
| Hispanic | 336 (38.4) | 317 (54.3) | 17 (7.8) | 2 (2.7) | |
| Declined | 31 (3.5) | 28 (4.8) | 2 (0.9) | 1 (1.4) | |
| Missing | 0 (0.0) | 0 (0.0) | 0 (0.0) | 0 (0.0) | |
| **Insurance** | | | | | **<0.001** |
| Private | 552 (63.1) | 281 (48.1) | 210 (96.3) | 61 (83.6) | |
| Medicaid | 321 (36.7) | 301 (51.5) | 8 (3.7) | 12 (16.4) | |
| Missing | 2 (0.2) | 2 (0.3) | 0 (0.0) | 0 (0.0) | |
| **Mode of delivery** | | | | | **0.015** |
| Vaginal | 559 (63.9) | 354 (60.6) | 159 (72.9) | 46 (63.0) | |
| C-Section | 316 (36.1) | 230 (39.4) | 59 (27.1) | 27 (37.0) | |
| Missing | 0 (0.0) | 0 (0.0) | 0 (0.0) | 0 (0.0) | |
| **Parity** | | | | | 0.588 |
| Multiparous | 472 (53.9) | 310 (53.1) | 117 (53.7) | 45 (61.6) | |
| Primiparous | 403 (46.1) | 274 (46.9) | 101 (46.3) | 28 (38.4) | |
| Missing | 0 (0.0) | 0 (0.0) | 0 (0.0) | 0 (0.0) | |
| **Infant sex** | | | | | |
| Male | 476 (53.8) | 334 (56.7) | 108 (48.9) | 34 (45.9) | 0.112 |
| Female | 408 (46.2) | 255 (43.3) | 113 (51.1) | 40 (54.1) | |
| Missing | 0 (0.0) | 0 (0.0) | 0 (0.0) | 0 (0.0) | |
| **Twin status** | | | | | |
| Singleton | 875 (99.0) | 584 (99.2) | 218 (98.6) | 73 (98.6) | 0.774 |
| Twin pairs | 9 (1.0) | 5 (0.8) | 3 (1.4) | 1 (1.4) | |
| Missing | 0 (0.0) | 0 (0.0) | 0 (0.0) | 0 (0.0) | |

| | | | | | |
|---|---|---|---|---|---|
| **Prematurity status** | | | | | |
| Not Preterm | 801 (90.6) | 523 (88.8) | 208 (94.1) | 70 (94.6) | 0.077 |
| Preterm | 83 (9.4) | 66 (11.2) | 13 (5.9) | 4 (5.4) | |
| Missing | 0 (0.0) | 0 (0.0) | 0 (0.0) | 0 (0.0) | |
| **Infant corrected age at survey completion** Mean (SD) | | | | | |
| Parenting confidence | 3.0 (2.0) | 2.3 (1.6) | 4.6 (1.7) | 4.6 (1.8) | **<0.001** |
| Bonding | 4.7 (1.6) | 4.2 (1.1) | 5.2 (1.8) | 5.3 (2.4) | **<0.001** |
| Parental stress | 6.0 (0.4) | 6.0 (0.4) | 5.9 (0.1) | 6.0 (0.1) | 0.262 |
| Infant socio-emotional development | 6.2 (0.9) | 6.1 (0.8) | 6.2 (1.0) | 6.5 (1.3) | **0.023** |
| **Infant corrected age at video visit** | | | | | |
| Mean (SD) | 5.4 (2.2) | 5.1 (1.4) | 5.7 (2.6) | 6.1 (3.2) | **0.006** |

**eTable3. Characteristics of mother-infant dyads by three SARS-CoV-2 exposure groups: negative, positive and positive – timing unknown**

| | Overall | Negative | Positive | Positive - Timing Unknown | *p*-value |
|---|---|---|---|---|---|
| | N=884 | n=568 | n=263 | n=53 | |
| **Maternal age at delivery** | | | | | |
| Mean (SD) | 32.2 (5.4) | 32.6 (5.3) | 31.8 (5.6) | 30.5 (5.2) | **0.02** |
| **Maternal race** | | | | | **<0.001** |
| White | 456 (52.1) | 334 (59.3) | 99 (38.1) | 23 (44.2) | |
| Other/More than one category | 180 (20.6) | 86 (15.3) | 84 (32.3) | 10 (19.2) | |
| Black/African-American | 100 (11.4) | 59 (10.5) | 36 (13.8) | 5 (9.6) | |
| Asian | 43 (4.9) | 36 (6.4) | 6 (2.3) | 1 (1.9) | |
| Native Hawaiian/ Pacific Islander | 7 (0.8) | 3 (0.5) | 3 (1.2) | 1 (1.9) | |
| Native American/ Alaska Native | 4 (0.5) | 3 (0.5) | 1 (0.4) | 0 (0.0) | |
| Declined | 84 (9.6) | 42 (7.5) | 30 (11.5) | 12 (23.1) | |
| Missing | 1 (0.1) | 0 (0.0) | 1 (0.4) | 0 (0.0) | |
| **Maternal ethnicity** | | | | | **<0.001** |
| Not Hispanic | 508 (58.1) | 380 (67.5) | 107 (41.2) | 21 (40.4) | |
| Hispanic | 336 (38.4) | 164 (29.1) | 142 (54.6) | 30 (57.7) | |
| Declined | 31 (3.5) | 19 (3.4) | 11 (4.2) | 1 (1.9) | |
| Missing | 0 (0.0) | 0 (0.0) | 0 (0.0) | 0 (0.0) | |
| **Insurance** | | | | | **<0.001** |
| Private | 552 (63.1) | 399 (70.9) | 133 (51.2) | 20 (38.5) | |
| Medicaid | 321 (36.7) | 163 (29.0) | 126 (48.5) | 32 (61.5) | |
| Missing | 2 (0.2) | 1 (0.2) | 1 (0.4) | 0 (0.0) | |
| **Mode of delivery** | | | | | 0.925 |
| Vaginal | 559 (63.9) | 355 (63.1) | 170 (65.4) | 34 (65.4) | |
| C-Section | 316 (36.1) | 208 (36.9) | 90 (34.6) | 18 (34.6) | |
| Missing | 0 (0.0) | 0 (0.0) | 0 (0.0) | 0 (0.0) | |
| **Parity** | | | | | 0.13 |
| Multiparous | 472 (53.9) | 287 (51.0) | 155 (59.6) | 30 (57.7) | |
| Primiparous | 403 (46.1) | 276 (49.0) | 105 (40.4) | 22 (42.3) | |
| Missing | 0 (0.0) | 0 (0.0) | 0 (0.0) | 0 (0.0) | |
| **Infant sex** | | | | | 0.992 |
| Male | 476 (53.8) | 308 (54.2) | 140 (53.2) | 28 (52.8) | |
| Female | 408 (46.2) | 260 (45.8) | 123 (46.8) | 25 (47.2) | |
| Missing | 0 (0.0) | 0 (0.0) | 0 (0.0) | 0 (0.0) | |
| **Twin sex** | | | | | 0.757 |
| Singleton | 866 (99.0) | 558 (99.1) | 257 (98.9) | 51 (98.1) | |
| Multiple | 9 (1.0) | 5 (0.9) | 3 (1.1) | 1 (1.9) | |

| | | | | | |
|---|---|---|---|---|---|
| Missing | 0 (0.0) | 0 (0.0) | 0 (0.0) | 0 (0.0) | |
| **Prematurity status** | | | | | 0.097 |
| Not Preterm | 801 (90.6) | 525 (92.4) | 229 (87.1) | 47 (88.7) | |
| Preterm | 83 (9.4) | 43 (7.6) | 34 (12.9) | 6 (11.3) | |
| Missing | 0 (0.0) | 0 (0.0) | 0 (0.0) | 0 (0.0) | |
| **Infant corrected age at survey completion** Mean (SD) | | | | | |
| Parenting confidence | 3.0 (2.0) | 3.0 (2.0) | 2.6 (1.8) | 4.5 (1.4) | **<0.001** |
| Bonding | 4.7 (1.6) | 4.7 (1.6) | 4.4 (1.4) | 5.0 (1.5) | 0.057 |
| Parental stress | 6.0 (0.4) | 6.0 (0.4) | 6.0 (0.4) | 6.0 (0.1) | 0.349 |
| Infant socio-emotional development | 6.2 (0.9) | 6.2 (0.9) | 6.1 (0.6) | 6.8 (1.6) | **<0.001** |
| **Infant corrected age at video visit** | | | | | |
| Mean (SD) | 5.4 (2.2) | 5.6 (2.3) | 4.8 (1.3) | 5.5 (2.7) | **0.027** |

**eTable4. Generalized linear model estimates of the effect of prenatal SARS-CoV-2 exposure on parenting stress (N=499)**

| | **Exponentiated estimate** | **Lower CI** | **Upper CI** | **p-value** |
|---|---|---|---|---|
| (Intercept) | 26.71 | 18.40 | 38.78 | <0.001 |
| SARS-CoV-2 prenatal exposure status: Negative | Reference | - | - | - |
| SARS-CoV-2 prenatal exposure status: Positive | 1.00 | 0.95 | 1.05 | 0.91 |
| Maternal age | 1.00 | 1.00 | 1.00 | 0.91 |
| Maternal race: White | Reference | - | - | - |
| Maternal race: Other | 0.97 | 0.91 | 1.02 | 0.24 |
| Maternal race: Black/African American | 0.91 | 0.84 | 0.98 | **0.01** |
| Maternal race: Declined | 0.96 | 0.87 | 1.06 | 0.39 |
| Maternal ethnicity: not Hispanic | Reference | - | - | - |
| Maternal ethnicity: Hispanic | 0.95 | 0.90 | 1.01 | 0.10 |
| Maternal ethnicity: Declined | 1.05 | 0.93 | 1.18 | 0.43 |
| Insurance: Commercial | Reference | - | - | - |
| Insurance: Medicaid | 0.96 | 0.91 | 1.02 | 0.18 |
| Mode of delivery: Vaginal | Reference | - | - | - |
| Mode of delivery: C-Section | 1.00 | 0.96 | 1.05 | 0.90 |
| Parity: Multiparous | Reference | - | - | - |
| Parity: Primiparous | 0.96 | 0.92 | 1.01 | 0.08 |
| Maternal mental health symptoms | 1.15 | 1.12 | 1.18 | **<0.001** |
| Infant sex: Male | Reference | - | - | - |
| Infant sex: Female | 0.99 | 0.95 | 1.04 | 0.75 |
| Infant corr. age at assessment | 1.06 | 1.00 | 1.12 | 0.05 |
| Study site: New York | Reference | - | - | - |
| Study site: Utah | 1.07 | 1.01 | 1.14 | **0.03** |
| Study site: Alabama | 0.98 | 0.87 | 1.11 | 0.80 |

**eTable5. Generalized linear model estimates of the effect of prenatal SARS-CoV-2 exposure on parenting confidence (N=621)**

| | Exponentiated estimate | Lower CI | Upper CI | p-value |
|---|---|---|---|---|
| (Intercept) | 37.24 | 33.61 | 41.26 | <0.001 |
| SARS-CoV-2 prenatal exposure status: Negative | Reference | - | - | - |
| SARS-CoV-2 prenatal exposure status: Positive | 1.01 | 0.98 | 1.03 | 0.61 |
| Maternal age | 1.00 | 1.00 | 1.00 | 0.56 |
| Maternal race: White | Reference | - | - | - |
| Maternal race: Other | 0.98 | 0.94 | 1.01 | 0.19 |
| Maternal race: Black/African American | 1.01 | 0.96 | 1.06 | 0.70 |
| Maternal race: Declined | 1.01 | 0.96 | 1.07 | 0.67 |
| Maternal ethnicity: not Hispanic | Reference | - | - | - |
| Maternal ethnicity: Hispanic | 1.02 | 0.99 | 1.06 | 0.21 |
| Maternal ethnicity: Declined | 1.02 | 0.94 | 1.10 | 0.69 |
| Insurance: Commercial | Reference | - | - | - |
| Insurance: Medicaid | 1.02 | 0.99 | 1.06 | 0.25 |
| Mode of delivery: Vaginal | Reference | - | - | - |
| Mode of delivery: C-Section | 1.00 | 0.98 | 1.03 | 0.78 |
| Parity: Multiparous | Reference | - | - | - |
| Parity: Primiparous | 0.98 | 0.95 | 1.01 | 0.11 |
| Maternal mental health symptoms | 0.96 | 0.94 | 0.97 | **<0.001** |
| Infant sex: Male | Reference | - | - | - |
| Infant sex: Female | 1.02 | 0.99 | 1.05 | 0.13 |
| Infant corr. age at assessment | 1.01 | 1.00 | 1.02 | **0.04** |
| Study site: New York | Reference | - | - | - |
| Study site: Utah | 1.03 | 0.99 | 1.08 | 0.19 |
| Study site: Alabama | 1.05 | 0.99 | 1.1025 | 0.09 |

**eTable6. Generalized linear model estimates of the effect of prenatal SARS-CoV-2 exposure on bonding (N=659)**

| | Exponentiated estimate | Lower CI | Upper CI | p-value |
|---|---|---|---|---|
| (Intercept) | 61.57 | 56.71 | 66.84 | <0.001 |
| SARS-CoV-2 prenatal exposure status: Negative | Reference | - | - | - |
| SARS-CoV-2 prenatal exposure status: Positive | 1.00 | 0.98 | 1.03 | 0.76 |
| Maternal age | 1.00 | 1.00 | 1.00 | 0.69 |
| Maternal race: White | Reference | - | - | - |
| Maternal race: Other | 1.01 | 0.98 | 1.04 | 0.55 |
| Maternal race: Black/African American | 1.03 | 1.00 | 1.07 | 0.05 |
| Maternal race: Declined | 1.02 | 0.97 | 1.06 | 0.49 |
| Maternal ethnicity: not Hispanic | Reference | - | - | - |
| Maternal ethnicity: Hispanic | 1.03 | 1.00 | 1.06 | 0.05 |
| Maternal ethnicity: Declined | 1.02 | 0.96 | 1.09 | 0.45 |
| Insurance: Commercial | Reference | - | - | - |
| Insurance: Medicaid | 1.03 | 1.00 | 1.06 | **0.03** |
| Mode of delivery: Vaginal | Reference | - | - | - |
| Mode of delivery: C-Section | 1.00 | 0.98 | 1.02 | 0.94 |
| Parity: Multiparous | Reference | - | - | - |
| Parity: Primiparous | 0.99 | 0.97 | 1.01 | 0.26 |
| Maternal mental health symptoms | 0.96 | 0.95 | 0.97 | **<0.001** |
| Infant sex: Male | Reference | - | - | - |
| Infant sex: Female | 1.00 | 0.99 | 1.02 | 0.67 |
| Infant corr. age at assessment | 1.00 | 0.99 | 1.01 | 0.55 |
| Study site: New York | Reference | - | - | - |
| Study site: Utah | 1.00 | 0.98 | 1.03 | 0.75 |
| Study site: Alabama | 1.03 | 0.99 | 1.07 | 0.14 |

**eTable7. Generalized linear model estimates of the effect of prenatal SARS-CoV-2 exposure on parental stress (sensitivity analysis: SARS-CoV-2 timing unknown removed from Positive group; N=487).**

| | Exponentiated estimate | Lower CI | Upper CI | p-value |
|---|---|---|---|---|
| (Intercept) | 27.25 | 18.75 | 39.59 | <0.001 |
| SARS-CoV-2 prenatal exposure status: Negative | Reference | - | - | - |
| SARS-CoV-2 prenatal exposure status: Positive | 1.00 | 0.96 | 1.05 | 0.95 |
| Maternal age | 1.00 | 1.00 | 1.00 | 0.73 |
| Maternal race: White | Reference | - | - | - |
| Maternal race: Other | 0.98 | 0.92 | 1.03 | 0.40 |
| Maternal race: Black/African American | 0.91 | 0.84 | 0.98 | **0.01** |
| Maternal race: Declined | 0.96 | 0.87 | 1.06 | 0.45 |
| Maternal ethnicity: not Hispanic | Reference | - | - | - |
| Maternal ethnicity: Hispanic | 0.95 | 0.90 | 1.01 | 0.12 |
| Maternal ethnicity: Declined | 1.04 | 0.93 | 1.17 | 0.47 |
| Insurance: Commercial | Reference | - | - | - |
| Insurance: Medicaid | 0.95 | 0.90 | 1.01 | 0.11 |
| Mode of delivery: Vaginal | Reference | - | - | - |
| Mode of delivery: C-Section | 1.01 | 0.96 | 1.05 | 0.85 |
| Parity: Multiparous | Reference | - | - | - |
| Parity: Primiparous | 0.95 | 0.91 | 1.00 | **0.04** |
| Maternal mental health symptoms | 1.15 | 1.12 | 1.18 | **<0.001** |
| Infant sex: Male | Reference | - | - | - |
| Infant sex: Female | 0.99 | 0.95 | 1.04 | 0.79 |
| Infant corr. age at assessment | 1.06 | 1.00 | 1.12 | 0.06 |
| Study site: New York | Reference | - | - | - |
| Study site: Utah | 1.07 | 1.01 | 1.14 | **0.03** |
| Study site: Alabama | 0.99 | 0.87 | 1.11 | 0.82 |

**eTable8. Generalized linear model estimates of the effect of prenatal SARS-CoV-2 exposure on <u>parenting confidence</u> (sensitivity analysis: SARS-CoV-2 timing unknown removed from Positive group; N=588).**

| | **Exponentiated estimate** | **Lower CI** | **Upper CI** | **p-value** |
|---|---|---|---|---|
| (Intercept) | 37.00 | 33.28 | 41.14 | <0.001 |
| SARS-CoV-2 prenatal exposure status: Negative | Reference | - | - | - |
| SARS-CoV-2 prenatal exposure status: Positive | 1.00 | 0.98 | 1.03 | 0.77 |
| Maternal age | 1.00 | 1.00 | 1.00 | 0.48 |
| Maternal race: White | Reference | - | - | - |
| Maternal race: Other | 0.98 | 0.94 | 1.01 | 0.22 |
| Maternal race: Black/African American | 1.01 | 0.97 | 1.06 | 0.60 |
| Maternal race: Declined | 1.01 | 0.96 | 1.07 | 0.61 |
| Maternal ethnicity: not Hispanic | Reference | - | - | - |
| Maternal ethnicity: Hispanic | 1.02 | 0.98 | 1.06 | 0.29 |
| Maternal ethnicity: Declined | 1.01 | 0.93 | 1.10 | 0.73 |
| Insurance: Commercial | Reference | - | - | - |
| Insurance: Medicaid | 1.02 | 0.99 | 1.06 | 0.23 |
| Mode of delivery: Vaginal | Reference | - | - | - |
| Mode of delivery: C-Section | 1.00 | 0.98 | 1.03 | 0.79 |
| Parity: Multiparous | Reference | - | - | - |
| Parity: Primiparous | 0.98 | 0.95 | 1.01 | 0.11 |
| Maternal mental health symptoms | 0.96 | 0.94 | 0.97 | **<0.001** |
| Infant sex: Male | Reference | - | - | - |
| Infant sex: Female | 1.02 | 0.99 | 1.05 | 0.14 |
| Infant corr. age at assessment | 1.01 | 1.00 | 1.02 | **0.05** |
| Study site: New York | Reference | - | - | - |
| Study site: Utah | 1.03 | 0.98 | 1.08 | 0.20 |
| Study site: Alabama | 1.05 | 0.99 | 1.10 | 0.10 |

**eTable9. Generalized linear model estimates of the effect of prenatal SARS-CoV-2 exposure on bonding (sensitivity analysis: SARS-CoV-2 timing unknown removed from Positive group; N=613).**

| | **Exponentiated estimate** | **Lower CI** | **Upper CI** | **p-value** |
|---|---|---|---|---|
| (Intercept) | 61.47 | 56.43 | 66.97 | <0.001 |
| SARS-CoV-2 prenatal exposure status: Negative | Reference | - | - | - |
| SARS-CoV-2 prenatal exposure status: Positive | 1.00 | 0.98 | 1.03 | 0.90 |
| Maternal age | 1.00 | 1.00 | 1.00 | 0.61 |
| Maternal race: White | Reference | - | - | - |
| Maternal race: Other | 1.01 | 0.98 | 1.04 | 0.46 |
| Maternal race: Black/African American | 1.05 | 1.01 | 1.08 | **0.01** |
| Maternal race: Declined | 1.02 | 0.97 | 1.07 | 0.42 |
| Maternal ethnicity: not Hispanic | Reference | - | - | - |
| Maternal ethnicity: Hispanic | 1.02 | 0.99 | 1.06 | 0.14 |
| Maternal ethnicity: Declined | 1.02 | 0.95 | 1.08 | 0.64 |
| Insurance: Commercial | Reference | - | - | - |
| Insurance: Medicaid | 1.03 | 1.00 | 1.06 | **0.03** |
| Mode of delivery: Vaginal | Reference | - | - | - |
| Mode of delivery: C-Section | 1.00 | 0.98 | 1.02 | 0.99 |
| Parity: Multiparous | Reference | - | - | - |
| Parity: Primiparous | 0.99 | 0.97 | 1.01 | 0.33 |
| Maternal mental health symptoms | 0.96 | 0.95 | 0.97 | **<0.001** |
| Infant sex: Male | Reference | - | - | - |
| Infant sex: Female | 1.00 | 0.98 | 1.02 | 0.88 |
| Infant corr. age at assessment | 1.00 | 0.99 | 1.00 | 0.40 |
| Study site: New York | Reference | - | - | - |
| Study site: Utah | 1.01 | 0.98 | 1.04 | 0.58 |
| Study site: Alabama | 1.03 | 0.99 | 1.07 | 0.14 |

**eTable10. Generalized linear model estimates of the effect of prenatal SARS-CoV-2 exposure on parental stress (sensitivity analysis: infants born in February 2020 removed from sample; N=496)**

| | **Exponentiated estimate** | **Lower CI** | **Upper CI** | **p-value** |
|---|---|---|---|---|
| (Intercept) | 26.73 | 18.39 | 38.85 | <0.001 |
| SARS-CoV-2 prenatal exposure status: Negative | Reference | - | - | - |
| SARS-CoV-2 prenatal exposure status: Positive | 1.00 | 0.95 | 1.04 | 0.89 |
| Maternal age | 1.00 | 1.00 | 1.00 | 0.89 |
| Maternal race: White | Reference | - | - | - |
| Maternal race: Other | 0.97 | 0.91 | 1.02 | 0.23 |
| Maternal race: Black/African American | 0.91 | 0.84 | 0.98 | **0.01** |
| Maternal race: Declined | 0.96 | 0.87 | 1.05 | 0.38 |
| Maternal ethnicity: not Hispanic | Reference | - | - | - |
| Maternal ethnicity: Hispanic | 0.95 | 0.89 | 1.01 | 0.10 |
| Maternal ethnicity: Declined | 1.05 | 0.93 | 1.18 | 0.44 |
| Insurance: Commercial | Reference | - | - | - |
| Insurance: Medicaid | 0.96 | 0.91 | 1.02 | 0.18 |
| Mode of delivery: Vaginal | Reference | - | - | - |
| Mode of delivery: C-Section | 1.00 | 0.96 | 1.05 | 0.86 |
| Parity: Multiparous | Reference | - | - | - |
| Parity: Primiparous | 0.96 | 0.92 | 1.01 | 0.08 |
| Maternal mental health symptoms | 1.15 | 1.12 | 1.18 | **<0.001** |
| Infant sex: Male | Reference | - | - | - |
| Infant sex: Female | 0.99 | 0.95 | 1.04 | 0.76 |
| Infant corr. age at assessment | 1.06 | 1.00 | 1.12 | 0.05 |
| Study site: New York | Reference | - | - | - |
| Study site: Utah | 1.07 | 1.00 | 1.14 | **0.04** |
| Study site: Alabama | 0.98 | 0.87 | 1.11 | 0.79 |

**eTable11. Generalized linear model estimates of the effect of prenatal SARS-CoV-2 exposure on parenting confidence (sensitivity analysis: infants born in February 2020 removed from sample; N=618)**

| | Exponentiated estimate | Lower CI | Upper CI | p-value |
|---|---|---|---|---|
| (Intercept) | 37.11 | 33.48 | 41.13 | <0.001 |
| SARS-CoV-2 prenatal exposure status: Negative | Reference | - | - | - |
| SARS-CoV-2 prenatal exposure status: Positive | 1.01 | 0.98 | 1.04 | 0.58 |
| Maternal age | 1.00 | 1.00 | 1.00 | 0.52 |
| Maternal race: White | Reference | - | - | - |
| Maternal race: Other | 0.98 | 0.94 | 1.01 | 0.21 |
| Maternal race: Black/African American | 1.01 | 0.97 | 1.06 | 0.67 |
| Maternal race: Declined | 1.01 | 0.96 | 1.07 | 0.64 |
| Maternal ethnicity: not Hispanic | Reference | - | - | - |
| Maternal ethnicity: Hispanic | 1.02 | 0.99 | 1.06 | 0.21 |
| Maternal ethnicity: Declined | 1.02 | 0.94 | 1.10 | 0.68 |
| Insurance: Commercial | Reference | - | - | - |
| Insurance: Medicaid | 1.02 | 0.99 | 1.06 | 0.23 |
| Mode of delivery: Vaginal | Reference | - | - | - |
| Mode of delivery: C-Section | 1.00 | 0.98 | 1.03 | 0.85 |
| Parity: Multiparous | Reference | - | - | - |
| Parity: Primiparous | 0.98 | 0.95 | 1.01 | 0.11 |
| Maternal mental health symptoms | 0.96 | 0.94 | 0.97 | **<0.001** |
| Infant sex: Male | Reference | - | - | - |
| Infant sex: Female | 1.02 | 0.99 | 1.05 | 0.13 |
| Infant corr. age at assessment | 1.01 | 1.00 | 1.02 | **0.04** |
| Study site: New York | Reference | - | - | - |
| Study site: Utah | 1.03 | 0.99 | 1.08 | 0.16 |
| Study site: Alabama | 1.05 | 1.00 | 1.11 | 0.08 |

**eTable12. Generalized linear model estimates of the effect of prenatal SARS-CoV-2 exposure on bonding (sensitivity analysis: infants born in February 2020 removed from sample; N=659)**

| | **Exponentiated estimate** | **Lower CI** | **Upper CI** | **p-value** |
|---|---|---|---|---|
| (Intercept) | 61.57 | 56.71 | 66.84 | <0.001 |
| SARS-CoV-2 prenatal exposure status: Negative | Reference | - | - | - |
| SARS-CoV-2 prenatal exposure status: Positive | 1.00 | 0.98 | 1.03 | 0.76 |
| Maternal age | 1.00 | 1.00 | 1.00 | 0.69 |
| Maternal race: White | Reference | - | - | - |
| Maternal race: Other | 1.01 | 0.98 | 1.04 | 0.55 |
| Maternal race: Black/African American | 1.03 | 1.00 | 1.07 | 0.05 |
| Maternal race: Declined | 1.02 | 0.97 | 1.06 | 0.49 |
| Maternal ethnicity: not Hispanic | Reference | - | - | - |
| Maternal ethnicity: Hispanic | 1.03 | 1.00 | 1.06 | 0.05 |
| Maternal ethnicity: Declined | 1.02 | 0.96 | 1.09 | 0.45 |
| Insurance: Commercial | Reference | - | - | - |
| Insurance: Medicaid | 1.03 | 1.00 | 1.06 | **0.03** |
| Mode of delivery: Vaginal | Reference | - | - | - |
| Mode of delivery: C-Section | 1.00 | 0.98 | 1.02 | 0.94 |
| Parity: Multiparous | Reference | - | - | - |
| Parity: Primiparous | 0.99 | 0.97 | 1.01 | 0.26 |
| Maternal mental health symptoms | 0.96 | 0.95 | 0.97 | **<0.001** |
| Infant sex: Male | Reference | - | - | - |
| Infant sex: Female | 1.00 | 0.99 | 1.02 | 0.67 |
| Infant corr. age at assessment | 1.00 | 0.99 | 1.01 | 0.55 |
| Study site: New York | Reference | - | - | - |
| Study site: Utah | 1.01 | 0.98 | 1.03 | 0.75 |
| Study site: Alabama | 1.03 | 0.99 | 1.07 | 0.14 |

**eTable13. Generalized linear model estimates of the effect of prenatal SARS-CoV-2 exposure on quality of maternal caregiving behaviors (N=317)**

| | **Exponentiated estimate** | **Lower CI** | **Upper CI** | **p-value** |
|---|---|---|---|---|
| (Intercept) | 28.39 | 23.39 | 34.45 | <0.001 |
| SARS-CoV-2 prenatal exposure status: Negative | Reference | - | - | - |
| SARS-CoV-2 prenatal exposure status: Positive | 0.95 | 0.90 | 1.00 | **0.03** |
| Maternal age | 1.01 | 1.00 | 1.01 | **0.002** |
| Maternal race: White | Reference | - | - | - |
| Maternal race: Other | 0.90 | 0.84 | 0.97 | **0.005** |
| Maternal race: Black/African American | 0.91 | 0.82 | 1.00 | 0.05 |
| Maternal race: Declined | 1.01 | 0.90 | 1.12 | 0.89 |
| Maternal ethnicity: not Hispanic | Reference | - | - | - |
| Maternal ethnicity: Hispanic | 1.01 | 0.94 | 1.09 | 0.76 |
| Maternal ethnicity: Declined | 1.06 | 0.91 | 1.25 | 0.45 |
| Insurance: Commercial | Reference | - | - | - |
| Insurance: Medicaid | 0.97 | 0.90 | 1.03 | 0.32 |
| Mode of delivery: Vaginal | Reference | - | - | - |
| Mode of delivery: C-Section | 0.99 | 0.94 | 1.03 | 0.52 |
| Parity: Multiparous | Reference | - | - | - |
| Parity: Primiparous | 1.012 | 0.97 | 1.07 | 0.46 |
| Maternal mental health symptoms | 1.01 | 0.99 | 1.04 | 0.36 |
| Infant sex: Male | Reference | - | - | - |
| Infant sex: Female | 1.02 | 0.98 | 1.07 | 0.34 |
| Infant corr. age at assessment | 0.99 | 0.98 | 1.01 | 0.26 |
| Study site: New York | Reference | - | - | - |
| Study site: Utah | 0.98 | 0.91 | 1.04 | 0.47 |
| Study site: Alabama | 0.91 | 0.84 | 1.00 | 0.05 |

**eTable14. Generalized linear model estimates of the effect of prenatal SARS-CoV-2 exposure on emotional connection (N=364)**

| | **Exponentiated estimate** | **Lower CI** | **Upper CI** | **p-value** |
|---|---|---|---|---|
| (Intercept) | 9.01 | 5.86 | 13.86 | <0.001 |
| SARS-CoV-2 prenatal exposure status: Negative | Reference | - | - | - |
| SARS-CoV-2 prenatal exposure status: Positive | 1.05 | 0.93 | 1.17 | 0.45 |
| Maternal age | 0.99 | 0.98 | 1.00 | 0.21 |
| Maternal race: White | Reference | - | - | - |
| Maternal race: Other | 0.94 | 0.80 | 1.10 | 0.43 |
| Maternal race: Black/African American | 1.04 | 0.85 | 1.26 | 0.73 |
| Maternal race: Declined | 0.77 | 0.61 | 0.97 | **0.03** |
| Maternal ethnicity: not Hispanic | Reference | - | - | - |
| Maternal ethnicity: Hispanic | 0.99 | 0.84 | 1.17 | 0.94 |
| Maternal ethnicity: Declined | 1.80 | 1.30 | 2.51 | **0.0004** |
| Insurance: Commercial | Reference | - | - | - |
| Insurance: Medicaid | 0.97 | 0.83 | 1.12 | 0.65 |
| Mode of delivery: Vaginal | Reference | - | - | - |
| Mode of delivery: C-Section | 0.98 | 0.87 | 1.09 | 0.65 |
| Parity: Multiparous | Reference | - | - | - |
| Parity: Primiparous | 0.84 | 0.76 | 0.94 | **0.002** |
| Maternal mental health symptoms | 1.00 | 0.95 | 1.05 | 0.99 |
| Infant sex: Male | Reference | - | - | - |
| Infant sex: Female | 0.95 | 0.8526 | 1.05 | 0.28 |
| Infant corr. age at assessment | 1.04 | 1.0107 | 1.06 | **0.004** |
| Study site: New York | Reference | - | - | - |
| Study site: Utah | 0.86 | 0.74 | 1.00 | 0.06 |
| Study site: Alabama | 0.80 | 0.65 | 0.97 | **0.03** |

**eTable15. Generalized linear model estimates of the effect of prenatal SARS-CoV-2 exposure on quality of maternal caregiving behaviors (sensitivity analysis: SARS-CoV-2 timing unknown removed from Positive group; N=291).**

| | Exponentiated estimate | Lower CI | Upper CI | p-value |
|---|---|---|---|---|
| (Intercept) | 29.68 | 24.41 | 36.09 | <0.001 |
| SARS-CoV-2 prenatal exposure status: Negative | Reference | - | - | - |
| SARS-CoV-2 prenatal exposure status: Positive | 0.95 | 0.90 | 1.00 | 0.05 |
| Maternal age | 1.01 | 1.00 | 1.01 | **0.01** |
| Maternal race: White | Reference | - | - | - |
| Maternal race: Other | 0.90 | 0.84 | 0.97 | **0.007** |
| Maternal race: Black/African American | 0.89 | 0.80 | 0.98 | **0.02** |
| Maternal race: Declined | 1.00 | 0.89 | 1.12 | 0.98 |
| Maternal ethnicity: not Hispanic | Reference | - | - | - |
| Maternal ethnicity: Hispanic | 1.00 | 0.92 | 1.08 | 0.94 |
| Maternal ethnicity: Declined | 1.09 | 0.93 | 1.29 | 0.28 |
| Insurance: Commercial | Reference | - | - | - |
| Insurance: Medicaid | 0.98 | 0.92 | 1.05 | 0.56 |
| Mode of delivery: Vaginal | Reference | - | - | - |
| Mode of delivery: C-Section | 0.98 | 0.93 | 1.03 | 0.48 |
| Parity: Multiparous | Reference | - | - | - |
| Parity: Primiparous | 1.02 | 0.97 | 1.07 | 0.39 |
| Maternal mental health symptoms | 1.02 | 0.99 | 1.04 | 0.17 |
| Infant sex: Male | Reference | - | - | - |
| Infant sex: Female | 1.03 | 0.99 | 1.09 | 0.10 |
| Infant corr. age at assessment | 0.99 | 0.98 | 1.00 | 0.16 |
| Study site: New York | Reference | - | - | - |
| Study site: Utah | 0.97 | 0.91 | 1.03 | 0.33 |
| Study site: Alabama | 0.93 | 0.85 | 1.01 | 0.09 |

**eTable16. Generalized linear model estimates of the effect of prenatal SARS-CoV-2 exposure on emotional connection (sensitivity analysis: SARS-CoV-2 timing unknown removed from Positive group; N=332).**

| | Exponentiated estimate | Lower CI | Upper CI | p-value |
|---|---|---|---|---|
| (Intercept) | 10.05 | 6.41 | 15.77 | <0.001 |
| SARS-CoV-2 prenatal exposure status: Negative | Reference | - | - | - |
| SARS-CoV-2 prenatal exposure status: Positive | 1.04 | 0.92 | 1.18 | 0.54 |
| Maternal age | 0.99 | 0.98 | 1.00 | 0.13 |
| Maternal race: White | Reference | - | - | - |
| Maternal race: Other | 0.92 | 0.78 | 1.09 | 0.34 |
| Maternal race: Black/African American | 1.04 | 0.85 | 1.28 | 0.70 |
| Maternal race: Declined | 0.80 | 0.63 | 1.02 | 0.07 |
| Maternal ethnicity: not Hispanic | Reference | - | - | - |
| Maternal ethnicity: Hispanic | 1.00 | 0.84 | 1.19 | 0.99 |
| Maternal ethnicity: Declined | 1.78 | 1.26 | 2.52 | **0.001** |
| Insurance: Commercial | Reference | - | - | - |
| Insurance: Medicaid | 0.96 | 0.82 | 1.12 | 0.57 |
| Mode of delivery: Vaginal | Reference | - | - | - |
| Mode of delivery: C-Section | 0.98 | 0.87 | 1.10 | 0.69 |
| Parity: Multiparous | Reference | - | - | - |
| Parity: Primiparous | 0.82 | 0.73 | 0.92 | **0.0008** |
| Maternal mental health symptoms | 1.02 | 0.96 | 1.07 | 0.59 |
| Infant sex: Male | Reference | - | - | - |
| Infant sex: Female | 0.94 | 0.85 | 1.05 | 0.27 |
| Infant corr. age at assessment | 1.03 | 1.01 | 1.06 | **0.02** |
| Study site: New York | Reference | - | - | - |
| Study site: Utah | 0.85 | 0.73 | 0.99 | **0.04** |
| Study site: Alabama | 0.79 | 0.64 | 0.96 | **0.02** |

**eTable17. Generalized linear model estimates of the effect of prenatal SARS-CoV-2 exposure on quality of maternal caregiving behaviors (sensitivity analysis: infants born in February 2020 removed from sample; N=317)**

| | Exponentiated estimate | Lower CI | Upper CI | p-value |
|---|---|---|---|---|
| (Intercept) | 28.39 | 23.39 | 34.45 | <0.001 |
| SARS-CoV-2 prenatal exposure status: Negative | Reference | - | - | - |
| SARS-CoV-2 prenatal exposure status: Positive | 0.95 | 0.90 | 1.00 | **0.03** |
| Maternal age | 1.01 | 1.00 | 1.01 | **0.002** |
| Maternal race: White | Reference | - | - | - |
| Maternal race: Other | 0.90 | 0.84 | 0.97 | **0.005** |
| Maternal race: Black/African American | 0.91 | 0.82 | 1.00 | 0.05 |
| Maternal race: Declined | 1.01 | 0.90 | 1.12 | 0.89 |
| Maternal ethnicity: not Hispanic | Reference | - | - | - |
| Maternal ethnicity: Hispanic | 1.01 | 0.94 | 1.09 | 0.76 |
| Maternal ethnicity: Declined | 1.06 | 0.91 | 1.25 | 0.45 |
| Insurance: Commercial | Reference | - | - | - |
| Insurance: Medicaid | 0.97 | 0.90 | 1.03 | 0.32 |
| Mode of delivery: Vaginal | Reference | - | - | - |
| Mode of delivery: C-Section | 0.98 | 0.94 | 1.03 | 0.52 |
| Parity: Multiparous | Reference | - | - | - |
| Parity: Primiparous | 1.02 | 0.97 | 1.07 | 0.46 |
| Maternal mental health symptoms | 1.01 | 0.99 | 1.04 | 0.36 |
| Infant sex: Male | Reference | - | - | - |
| Infant sex: Female | 1.02 | 0.98 | 1.07 | 0.34 |
| Infant corr. age at assessment | 0.99 | 0.98 | 1.01 | 0.26 |
| Study site: New York | Reference | - | - | - |
| Study site: Utah | 0.98 | 0.91 | 1.04 | 0.47 |
| Study site: Alabama | 0.91 | 0.84 | 1.00 | 0.05 |

**eTable18. Generalized linear model estimates of the effect of prenatal SARS-CoV-2 exposure on emotional connection (sensitivity analysis: infants born in February 2020 removed from sample; N=364)**

| | Exponentiated estimate | Lower CI | Upper CI | p-value |
|---|---|---|---|---|
| (Intercept) | 9.01 | 5.86 | 13.86 | <0.001 |
| SARS-CoV-2 prenatal exposure status: Negative | Reference | - | - | - |
| SARS-CoV-2 prenatal exposure status: Positive | 1.05 | 0.93 | 1.17 | 0.45 |
| Maternal age | 0.99 | 0.98 | 1.00 | 0.22 |
| Maternal race: White | Reference | - | - | - |
| Maternal race: Other | 0.94 | 0.80 | 1.10 | 0.43 |
| Maternal race: Black/African American | 1.04 | 0.85 | 1.26 | 0.73 |
| Maternal race: Declined | 0.77 | 0.61 | 0.97 | **0.03** |
| Maternal ethnicity: not Hispanic | Reference | - | - | - |
| Maternal ethnicity: Hispanic | 0.99 | 0.84 | 1.17 | 0.94 |
| Maternal ethnicity: Declined | 1.80 | 1.30 | 2.51 | **0.0004** |
| Insurance: Commercial | Reference | - | - | - |
| Insurance: Medicaid | 0.97 | 0.83 | 1.12 | 0.65 |
| Mode of delivery: Vaginal | Reference | - | - | - |
| Mode of delivery: C-Section | 0.98 | 0.87 | 1.09 | 0.65 |
| Parity: Multiparous | Reference | - | - | - |
| Parity: Primiparous | 0.84 | 0.76 | 0.94 | **0.002** |
| Maternal mental health symptoms | 1.00 | 0.95 | 1.05 | 0.99 |
| Infant sex: Male | Reference | - | - | - |
| Infant sex: Female | 0.95 | 0.85 | 1.05 | 0.28 |
| Infant corr. age at assessment | 1.04 | 1.01 | 1.06 | **0.004** |
| Study site: New York | Reference | - | - | - |
| Study site: Utah | 0.86 | 0.74 | 1.00 | 0.06 |
| Study site: Alabama | 0.80 | 0.65 | 0.97 | **0.03** |

**eTable19. Generalized linear model estimates of the effect of prenatal SARS-CoV-2 exposure on socio-emotional functioning (N=696)**

| | Exponentiated estimate | Lower CI | Upper CI | p-value |
|---|---|---|---|---|
| (Intercept) | 22.85 | 10.24 | 50.50 | <0.001 |
| SARS-CoV-2 prenatal exposure status: Negative | Reference | - | - | - |
| SARS-CoV-2 prenatal exposure status: Positive | 1.06 | 0.93 | 1.20 | 0.41 |
| Maternal age | 0.99 | 0.98 | 1.00 | 0.18 |
| Maternal race: White | Reference | - | - | - |
| Maternal race: Other | 1.04 | 0.88 | 1.23 | 0.64 |
| Maternal race: Black/African American | 0.93 | 0.75 | 1.15 | 0.47 |
| Maternal race: Declined | 1.04 | 0.80 | 1.36 | 0.77 |
| Maternal ethnicity: not Hispanic | Reference | - | - | - |
| Maternal ethnicity: Hispanic | 1.05 | 0.88 | 1.26 | 0.60 |
| Maternal ethnicity: Declined | 1.10 | 0.79 | 1.57 | 0.57 |
| Insurance: Commercial | Reference | - | - | - |
| Insurance: Medicaid | 1.17 | 0.99 | 1.39 | 0.06 |
| Mode of delivery: Vaginal | Reference | - | - | - |
| Mode of delivery: C-Section | 1.02 | 0.90 | 1.15 | 0.80 |
| Parity: Multiparous | Reference | - | - | - |
| Parity: Primiparous | 1.16 | 1.02 | 1.31 | **0.02** |
| Maternal mental health symptoms | 1.21 | 1.14 | 1.29 | **<0.001** |
| Infant sex: Male | Reference | - | - | - |
| Infant sex: Female | 1.06 | 0.94 | 1.19 | 0.34 |
| Infant corr. age at assessment | 1.00 | 0.90 | 1.12 | 0.95 |
| Study site: New York | Reference | - | - | - |
| Study site: Utah | 0.94 | 0.80 | 1.12 | 0.49 |
| Study site: Alabama | 0.92 | 0.73 | 1.16 | 0.46 |

**eTable20. Generalized linear model estimates of the effect of prenatal SARS-CoV-2 exposure on socio-emotional functioning risk (N=696)**

| | Exponentiated estimate | Lower CI | Upper CI | p-value |
|---|---|---|---|---|
| (Intercept) | 0.31 | 0.01 | 12.47 | 0.52 |
| SARS-CoV-2 prenatal exposure status: Negative | Reference | - | - | - |
| SARS-CoV-2 prenatal exposure status: Positive | 1.19 | 0.67 | 2.08 | 0.55 |
| Maternal age | 0.94 | 0.89 | 1.00 | **0.04** |
| Maternal race: White | Reference | - | - | - |
| Maternal race: Other | 0.99 | 0.48 | 1.99 | 0.97 |
| Maternal race: Black/African American | 0.56 | 0.21 | 1.38 | 0.23 |
| Maternal race: Declined | 0.71 | 0.21 | 2.11 | 0.55 |
| Maternal ethnicity: not Hispanic | Reference | - | - | - |
| Maternal ethnicity: Hispanic | 1.05 | 0.48 | 2.30 | 0.91 |
| Maternal ethnicity: Declined | 1.21 | 0.28 | 4.47 | 0.79 |
| Insurance: Commercial | Reference | - | - | - |
| Insurance: Medicaid | 1.71 | 0.85 | 3.48 | 0.13 |
| Mode of delivery: Vaginal | Reference | - | - | - |
| Mode of delivery: C-Section | 1.50 | 0.87 | 2.58 | 0.14 |
| Parity: Multiparous | Reference | - | - | - |
| Parity: Primiparous | 2.04 | 1.16 | 3.64 | **0.02** |
| Maternal mental health symptoms | 2.01 | 1.59 | 2.57 | **<0.001** |
| Infant sex: Male | Reference | - | - | - |
| Infant sex: Female | 1.32 | 0.78 | 2.23 | 0.30 |
| Infant corr. age at assessment | 1.01 | 0.59 | 1.66 | 0.98 |
| Study site: New York | Reference | - | - | - |
| Study site: Utah | 0.42 | 0.17 | 0.95 | **0.04** |
| Study site: Alabama | 0.85 | 0.27 | 2.35 | 0.77 |

**Note.** Risk is determined with cut-off score on the Ages and Stages Questionnaire: Socio-Emotional-2 as a binary outcome.

**eTable21. Generalized linear model estimates of the effect of prenatal SARS-CoV-2 exposure on infant socio-emotional functioning (sensitivity analysis: SARS-CoV-2 timing unknown removed from Positive group; N=661).**

| | **Exponentiated estimate** | **Lower CI** | **Upper CI** | **p-value** |
|---|---|---|---|---|
| (Intercept) | 23.00 | 9.34 | 55.83 | <0.001 |
| SARS-CoV-2 prenatal exposure status: Negative | Reference | - | - | - |
| SARS-CoV-2 prenatal exposure status: Positive | 1.03 | 0.90 | 1.18 | 0.66 |
| Maternal age | 0.99 | 0.98 | 1.01 | 0.21 |
| Maternal race: White | Reference | - | - | - |
| Maternal race: Other | 1.05 | 0.89 | 1.25 | 0.56 |
| Maternal race: Black/African American | 0.93 | 0.75 | 1.16 | 0.51 |
| Maternal race: Declined | 1.09 | 0.83 | 1.45 | 0.53 |
| Maternal ethnicity: not Hispanic | Reference | - | - | - |
| Maternal ethnicity: Hispanic | 1.05 | 0.87 | 1.27 | 0.59 |
| Maternal ethnicity: Declined | 1.11 | 0.79 | 1.60 | 0.56 |
| Insurance: Commercial | Reference | - | - | - |
| Insurance: Medicaid | 1.15 | 0.96 | 1.38 | 0.11 |
| Mode of delivery: Vaginal | Reference | - | - | - |
| Mode of delivery: C-Section | 1.01 | 0.89 | 1.15 | 0.85 |
| Parity: Multiparous | Reference | - | - | - |
| Parity: Primiparous | 1.14 | 1.00 | 1.31 | 0.05 |
| Maternal mental health symptoms | 1.23 | 1.15 | 1.32 | **<0.001** |
| Infant sex: Male | Reference | - | - | - |
| Infant sex: Female | 1.08 | 0.95 | 1.22 | 0.22 |
| Infant corr. age at assessment | 1.00 | 0.88 | 1.14 | 0.99 |
| Study site: New York | Reference | - | - | - |
| Study site: Utah | 0.94 | 0.79 | 1.11 | 0.44 |
| Study site: Alabama | 0.92 | 0.73 | 1.18 | 0.50 |

**eTable22. Generalized linear model estimates of the effect of prenatal SARS-CoV-2 exposure on infant socio-emotional functioning risk (sensitivity analysis: SARS-CoV-2 timing unknown removed from Positive group; N=661)**

| | **Exponentiated estimate** | **Lower CI** | **Upper CI** | **p-value** |
|---|---|---|---|---|
| (Intercept) | 0.09 | 0.00 | 4.00 | 0.20 |
| SARS-CoV-2 prenatal exposure status: Negative | Reference | - | - | - |
| SARS-CoV-2 prenatal exposure status: Positive | 1.12 | 0.61 | 2.01 | 0.71 |
| Maternal age | 0.95 | 0.90 | 1.00 | 0.07 |
| Maternal race: White | Reference | - | - | - |
| Maternal race: Other | 1.01 | 0.48 | 2.10 | 0.98 |
| Maternal race: Black/African American | 0.52 | 0.18 | 1.34 | 0.19 |
| Maternal race: Declined | 0.84 | 0.25 | 2.60 | 0.77 |
| Maternal ethnicity: not Hispanic | Reference | - | - | - |
| Maternal ethnicity: Hispanic | 1.03 | 0.45 | 2.34 | 0.94 |
| Maternal ethnicity: Declined | 1.41 | 0.31 | 5.34 | 0.63 |
| Insurance: Commercial | Reference | - | - | - |
| Insurance: Medicaid | 1.74 | 0.85 | 3.62 | 0.13 |
| Mode of delivery: Vaginal | Reference | - | - | - |
| Mode of delivery: C-Section | 1.52 | 0.86 | 2.65 | 0.15 |
| Parity: Multiparous | Reference | - | - | - |
| Parity: Primiparous | 1.79 | 0.99 | 3.27 | 0.06 |
| Maternal mental health symptoms | 2.09 | 1.63 | 2.70 | **<0.001** |
| Infant sex: Male | Reference | - | - | - |
| Infant sex: Female | 1.44 | 0.84 | 2.49 | 0.19 |
| Infant corr. age at assessment | 1.20 | 0.69 | 1.98 | 0.49 |
| Study site: New York | Reference | - | - | - |
| Study site: Utah | 0.40 | 0.16 | 0.93 | **0.04** |
| Study site: Alabama | 0.88 | 0.28 | 2.46 | 0.82 |

**Note.** Risk is determined with cut-off score on the Ages and Stages Questionnaire: Socio-Emotional-2.

**eTable23. Generalized linear model estimates of the effect of prenatal SARS-CoV-2 exposure on infant socio-emotional functioning (sensitivity analysis: infants born in February 2020 removed from sample; N=693)**

| | Exponentiated estimate | Lower CI | Upper CI | p-value |
|---|---|---|---|---|
| (Intercept) | 23.20 | 10.43 | 51.15 | <0.001 |
| SARS-CoV-2 prenatal exposure status: Negative | Reference | - | - | - |
| SARS-CoV-2 prenatal exposure status: Positive | 1.06 | 0.93 | 1.20 | 0.41 |
| Maternal age | 0.99 | 0.98 | 1.00 | 0.15 |
| Maternal race: White | Reference | - | - | - |
| Maternal race: Other | 1.04 | 0.88 | 1.23 | 0.62 |
| Maternal race: Black/African American | 0.93 | 0.75 | 1.14 | 0.47 |
| Maternal race: Declined | 1.05 | 0.81 | 1.37 | 0.74 |
| Maternal ethnicity: not Hispanic | Reference | - | - | - |
| Maternal ethnicity: Hispanic | 1.04 | 0.87 | 1.25 | 0.65 |
| Maternal ethnicity: Declined | 1.10 | 0.79 | 1.56 | 0.59 |
| Insurance: Commercial | Reference | - | - | - |
| Insurance: Medicaid | 1.16 | 0.98 | 1.38 | 0.07 |
| Mode of delivery: Vaginal | Reference | - | - | - |
| Mode of delivery: C-Section | 1.02 | 0.90 | 1.16 | 0.78 |
| Parity: Multiparous | Reference | - | - | - |
| Parity: Primiparous | 1.16 | 1.02 | 1.31 | **0.03** |
| Maternal mental health symptoms | 1.21 | 1.14 | 1.29 | **<0.001** |
| Infant sex: Male | Reference | - | - | - |
| Infant sex: Female | 1.05 | 0.94 | 1.19 | 0.39 |
| Infant corr. age at assessment | 1.01 | 0.90 | 1.13 | 0.93 |
| Study site: New York | Reference | - | - | - |
| Study site: Utah | 0.94 | 0.79 | 1.11 | 0.45 |
| Study site: Alabama | 0.91 | 0.73 | 1.16 | 0.44 |

**eTable24. Generalized linear model estimates of the effect of prenatal SARS-CoV-2 exposure on infant socio-emotional functioning risk (sensitivity analysis: infants born in February 2020 removed from sample; N=693)**

| | **Exponentiated estimate** | **Lower CI** | **Upper CI** | **p-value** |
|---|---|---|---|---|
| (Intercept) | 0.34 | 0.01 | 13.79 | 0.55 |
| SARS-CoV-2 prenatal exposure status: Negative | Reference | - | - | - |
| SARS-CoV-2 prenatal exposure status: Positive | 1.22 | 0.69 | 2.14 | 0.49 |
| Maternal age | 0.94 | 0.89 | 0.99 | **0.03** |
| Maternal race: White | Reference | - | - | - |
| Maternal race: Other | 1.05 | 0.51 | 2.13 | 0.90 |
| Maternal race: Black/African American | 0.59 | 0.22 | 1.45 | 0.27 |
| Maternal race: Declined | 0.77 | 0.23 | 2.30 | 0.65 |
| Maternal ethnicity: not Hispanic | Reference | - | - | - |
| Maternal ethnicity: Hispanic | 0.98 | 0.44 | 2.15 | 0.95 |
| Maternal ethnicity: Declined | 1.15 | 0.26 | 4.24 | 0.84 |
| Insurance: Commercial | Reference | - | - | - |
| Insurance: Medicaid | 1.63 | 0.80 | 3.31 | 0.18 |
| Mode of delivery: Vaginal | Reference | - | - | - |
| Mode of delivery: C-Section | 1.48 | 0.86 | 2.55 | 0.16 |
| Parity: Primiparous | 1.96 | 1.11 | 3.51 | 0.02 |
| Maternal mental health symptoms | 2.01 | 1.58 | 2.57 | **<0.001** |
| Infant sex: Male | Reference | - | - | - |
| Infant sex: Female | 1.28 | 0.75 | 2.17 | 0.36 |
| Infant corr. age at assessment | 1.02 | 0.59 | 1.68 | 0.94 |
| Study site: New York | Reference | - | - | - |
| Study site: Utah | 0.41 | 0.17 | 0.94 | **0.04** |
| Study site: Alabama | 0.84 | 0.27 | 2.31 | 0.74 |

**Note.** Risk is determined with cut-off score on the Ages and Stages Questionnaire: Socio-Emotional-2.
